# The effects of a 20-week exercise program on blood-circulating biomarkers related to brain health in children with overweight or obesity: The ActiveBrains project

**DOI:** 10.1101/2022.07.28.22278146

**Authors:** María Rodriguez-Ayllon, Abel Plaza-Florido, Andrea Mendez-Gutierrez, Signe Altmäe, Patricio Solis- Urra, Concepción M. Aguilera, Andrés Catena, Francisco B. Ortega, Irene Esteban-Cornejo

**Author notes:** **Corresponding author:** Francisco B Ortega, Department of Physical Education and Sports, Faculty of Sport Sciences, University of Granada (Spain), Carretera de Alfacar, S/N C.P. 18071 (Granada) Granada.

## Abstract

**Background:** Emerging research supports that exercise positively affects neurodevelopment. However, the mechanisms linking exercise with brain health are largely unknown. We aimed to investigate the effect of exercise on (i) blood biomarkers selected based on previous evidence (brain-derived neurotrophic factor (BDNF), β-hydroxybutyrate (BHB), cathepsin B (CTSB), kynurenine, fibroblast growth factor 21 (FGF21), vascular cell adhesion molecule-1 (sVCAM-1)); and (ii) a panel of 92 neurology-related proteins (discovery analysis). We also investigated whether changes in these biomarkers mediate the effects of exercise on brain health (hippocampal structure and function, cognitive performance, and mental health).

**Methods:** 81 children with overweight/obesity (10.1 ±1.1 years, 41% girls) were randomized to either 20-week of aerobic plus resistance exercise or control. Candidate biomarkers were assessed using ELISA for Kynurenine, FGF21, and CTSB, colorimetry for BHB, and XMap for BDNF and VCAM-1. The 92 neurology-related proteins were analyzed by antibody-based proteomic analyses.

**Results:** Our intervention had no significant effect on candidate biomarkers (all p>0.05). In the discovery analysis, a reduction in circulating macrophage scavenger receptor type-I (MSR1) was observed (standardized differences between groups (SMD): -0.3, p=0.001). This effect was validated using ELISA methods (SMD: -0.3, p=0.01). None of the biomarkers mediated the effects of exercise on brain health.

**Conclusions:** Our study does not support a chronic effect of exercise on candidate biomarkers. Nevertheless, we observed that chronic exercise reduced the levels of MRS1, while it did not mediate the effects of exercise on brain health. Future studies should explore the implications of this novel biomarker on general health.

**Highlights:**

- Candidate biomarkers (i.e., brain-derived neurotrophic factor (BDNF), β-hydroxybutyrate (BHB), cathepsin B (CTSB), kynurenine, fibroblast growth factor 21 (FGF21), vascular cell adhesion molecule-1 (sVCAM-1)) did not mediate the effects of exercise on brain health outcomes.
- Our discovery analysis, a panel of 92 neurology-related proteins, identified a reduction effect of exercise on blood-circulating MRS1.
- Exercise-induced changes in MRS1 did not mediate changes in brain health outcomes.
- The MSR1 is a membrane glycoprotein, that has not been related to exercise before, expressed in macrophages, and involved in pathological processes such as atherosclerosis, non-alcoholic fatty liver disease, and neurological diseases.
- Future studies should confirm the effect of exercise on MRS1 and its implications on general health.

## 1. Introduction

Childhood obesity is a major health concern, negatively associated with brain health ^1–3^. Exercise positively affects neurodevelopment during childhood ^4–6^, which in turn might protect the brain against the adverse effects of obesity at early stages in life. However, molecular mechanisms linking exercise with brain health indicators are still lacking. In this regard, brain-derived neurotrophic factor (BDNF) is a neurotrophin involved in the growth and healthy maintenance of neurons and exerts its major activity within the central nervous system ^7–9^. A growing number of studies have indicated that decreased levels of circulating BDNF are associated with cognitive decline and reduced performance on learning and memory tasks ^10^ as well as with anxiety and depression in animals and adult humans ^11,12^.

Exercise produces chronic benefits in the brain that enhance cognitive function and mental health ^13^. For instance, chronic exercise leads to an increase in BDNF in the central nervous system to promote improvement in cognitive ability and anxiety and depressive-like behaviors in animal models ^14–16^. In adults, previous exercise interventions showed improvement in hippocampal plasticity and memory, associated to BDNF changes ^17,18^. Though these results are widely recognized, the molecular mechanisms that elicit these positive effects of chronic exercise on brain health indicators (i.e., BDNF, brain structure, brain function, cognition, and mental health) in a period of life when brain is growing and developing, i.e. childhood, are still lacking.

Due to ethical considerations, blood is the primary tissue to study the molecular response to exercise in children, being muscle biopsy for instance rarely used in children for research purposes ^19^. In this regard, recent studies in animal models have tested several hypotheses to identify novel circulating biomarkers influenced by exercise ^20–22^, which could be helpful to understand how chronic exercise induces changes in different brain health indicators ^23^. First, the metabolite β-hydroxybutyrate (BHB) appears to increase chronically in the liver after exercise intervention ^21^. Particularly, it crosses the blood-brain barrier (BBB) and activates *BDNF* promoters in the hippocampus. This induction, in turn, mediates chronic exercise’s positive effects on memory, cognition, and synaptic transmission ^21^. Second, cathepsin B (CTSB) seems to cross the BBB, and increases BDNF levels ^23^. As such, this could affect hippocampal neurogenesis, hippocampal-dependent learning, memory, and depression ^22,23^. Third, exercise increases the PGC1α-dependent muscular expression of kynurenine aminotransferase enzymes ^24^. Likewise, higher expression of kynurenine aminotransferase induces a beneficial shift in the balance between the neurotoxic kynurenine and the neuroprotective kynurenic acid ^25^. The fact that kynurenic acid is not able to cross the BBB protects the brain from stress-induced kynurenine accumulation, neuroinflammation, changes in synaptic plasticity, and depressive symptoms ^24–26^. Fourth, plasma fibroblast growth factor 21 (FGF21) can cross the BBB ^27^, decrease oxidative stress and neuroinflammation, and in turn improve cognition ^28,29^. Meanwhile, a positive association between exercise and circulating FGF21 levels has been reported in some, but not all studies in older people ^29^. Lastly, elevated soluble vascular cell adhesion molecule-1 (sVCAM-1) seems to promote neuroinflammation ^30,31^, which can have negative consequences on cognitive function and emotional regulation ^30^. Although more research is needed to confirm this, literature suggests exercise could reduce sVCAM-1 in children ^32^. Collectively, despite these interesting observations, the effect of chronic exercise on BDNF, BHB, CTSB, Kynurenine, FGF21, and sVCAM-1, as well as its mediating role on brain health during childhood remain unexplored.

Besides target molecules, the effects of exercise on brain health could also be related to a more complex group of proteins and their interactions. Therefore, we need to use a more comprehensive set of plasma proteins to understand such complex patterns. In this context, high-throughput proteomic analysis methods permit the quantification of several proteins in plasma simultaneously ^33^, which allows exploring a broad set of blood-circulating biomarkers related to the beneficial effects of exercise on brain health during childhood. Therefore, the primary aims of this study are: (i) to examine whether a 20-week exercise program has an effect on selected blood-circulating biomarkers in children with OW/OB (candidate analyses); and (i) to explore the effect of a 20-week exercise program on 92 neurology-related plasma proteins, which may reveal new candidate biomarkers related to exercise (discovery analyses). As a secondary aim, we investigate whether changes in these emerging blood-circulating biomarkers, mediate the effects of exercise on brain health (i.e., hippocampal structure and function, cognition, and mental health) in children with OW/OB. As we exposed above, some blood-circulating biomarkers are expected to activate BDNF. Therefore, we will also explore whether changes in some blood-circulating biomarkers mediate the effect of exercise on BDNF levels. Overall, our main hypothesis was that chronic exercise induces changes in circulating blood levels of the selected biomarkers, which may induce a beneficial effect on brain health during childhood.

## 2. Methods

### 2.1 Design and participants

Children from the ActiveBrains project (http://profith.ugr.es/activebrains) participated in this study. It is a randomized controlled trial (RCT) designed to examine the effects of a 20-weeks physical exercise program on brain, cognitive and academic performance, as well as on selected physical and mental health outcomes in children with OW/OB. The ActiveBrains project was approved by the Human Research Ethics Committee of the University of Granada. Additionally, it was registered in ClinicalTrials.gov (identifier: NCT02295072).

Details of the ActiveBrains project methodology have been described elsewhere ^34^. Briefly, the eligibility criteria were: i) to be 8 to 11.9 years-old; ii) to have OW/OB according to the World Obesity Federation cut-off points ^35,36^; iii) not to suffer from physical disabilities or neurological disorders that impeded them to exercise; iv) in the case of girls, not to have started the menstruation at the moment of baseline assessments; v) to report no use of medications that influenced central nervous system function; vi) to be right-handed (i.e., measured by the Edinburgh inventory) ^37^; and vii) not to report an attention-deficit hyperactivity disorder (ADHD) over the 85th percentile measured by the ADHD rating scale ^38^.

Further details related to the randomization, physical exercise intervention, and control group condition have been described by Ortega et al. ^39^.

### 2.2 Blood-circulating biomarkers

Blood samples were obtained for biochemical and hematological screening tests between 8.30 AM and 10.30 AM. The blood collected into tubes containing EDTA was centrifuged (10 min at 4°C, 1000g). Plasma was isolated aliquoted and stored at -80° C for further analysis.

#### 2.2.1 Candidate blood-circulating biomarkers

First, the analysis of mature BDNF levels in plasma was performed using XMap technology (Luminex Corporation, Austin, TX), and using human monoclonal antibodies (Milliplex Map Kit, Millipore, Billerica, MA). For mature BDNF, we used the Human Neurodegenerative Disease Magnetic Bead Panel 3 (catalog HNDG3MAG-36K; EMD Millipore Corporation, Billerica, MA) with a sensitivity of 0.23 pg/mL, and intra- and inter-assay precision CV of <5.4% and <5.3%, respectively. Second, plasma kynurenine levels were quantified by enzyme-linked immune-absorbent assay (ELISA) (BA E-2200, LDN, Nordhorn, Germany, coefficient of variance (CV)=12.9), according to the manufacturer’s instructions. Second, serum levels of FGF21 and CTSB were measured by ELISA, using the kits RD191108200R (Biovendor, Brno, Czech Republic, CV=7.62) and ab272205 (Abcam, Cambridge, United Kingdom (UK), CV=2.91), respectively. Third, serum BHB concentration was quantified by a colorimetric assay (ab83390, Abcam, Cambridge, UK, CV=4.27) and sVCAM-1 was determined with XMap technology (Luminex Corporation, Austin, TX) using HNDG3MAG-36K (Millipore, Billerica, MA, United States of America (USA), CV=3.36).

#### 2.2.2 Panel of neurology-related proteins

The 92 neurology-related proteins were analyzed at the Olink laboratory in Uppsala by the Proximity Extension Assay technique using the Proseek Multiplex Neurology I 96 × 96 reagents kit (Olink Bioscience, Uppsala, Sweden), as previously described ^40,41^. The proteins included in the Neurology panel has been listed in **Table S1A**. Data are presented as arbitrary units and particularly as NPX (Normalized Protein eXpression). In brief, NPX is the Olink’s arbitrary unit which is in log2 scale, where high NPX value equals a high protein concentration. Intra- and inter-assay CV, detection limits, and specific information for each protein are described on the manufacturer’s website (https://www.olink.com/). The neurology panel includes a mix of proteins related to neurobiological processes and neurological diseases (e.g. neural development, axon guidance or synaptic function), as well as some more exploratory proteins with broader roles in processes such as cellular regulation, immunology, development and metabolism (https://www.olink.com/products/neurology/). From the 92 proteins included in the Olink proteomic neurology panel (https://www.olink.com/products/neurology/), a total of 91 proteins (99%) were detected in our sample, while Microtubule-associated protein tau (MAPT) did not pass the detection control criteria (proteins detected in >75% of the sample).

The effect of exercise on some of the proteins assessed by the Proximity Extension Assay technique was validated with ELISA. Particularly, we validated Kynureninase (KYNU), macrophage scavenger receptor type-I (MSR1), and plexin-B3 (PLXNB3), using the kits ELH-KYNU-1 (RayBiotech, Norcross, GA, USA, CV=12.4) SK00489-01 (Aviscera Bioscience, Santa Clara, CA, USA, CV=8.25) and EK1827 (Boster Bio, Pleasanton, CA, USA, CV=7.71), respectively. The decision of validating these specific neurology-related proteins (i.e., KYNU, MSR1, and PLXNB3) were based on (i) the effect size observed in the main analyses; and/or (iii) their relationship with BDNF (see **Table S2A**).

### 2.3 Outcome Measurements

Details related to the hippocampal volumetric analyses, hippocampal functional connectivity analysis, cognitive performance, and mental health measurements have been previously described ^39,42–44^.

### 2.4 Statistical analysis

#### 2.4.1 Main analyses: Effects of chronic exercise on circulating neurology-related proteins

Statistical procedures were performed using the SPSS software (version 22.0, IBM Corporation) and R version 1.3.1073. A significance difference level of p *<* 0.05 was set. Characteristics of the study sample are presented as mean and standard deviation (SD), or frequency and percentage as appropriate.

Main effects on the blood-circulating biomarkers were tested according to the per-protocol principle (participants with valid data who attended at least 70% of the recommended 3 sessions/week). Results from the per-protocol analysis were reported as the main findings as we were interested in knowing the efficacy rather than the effectiveness of our exercise intervention, i.e. we wanted to know the effects of exercise on the study biomarkers when exercise was actually done (operationally defined as ≥70% attendance). Main analyses were performed with analysis of covariance (ANCOVA) using post-exercise program data as dependent variables, group (i.e., exercise vs. control) as fixed factor, and baseline data as covariates. The z-scores for each blood-circulating biomarkers at post-exercise program were formed by dividing the difference of the raw score of each participant from the baseline mean by the baseline standard deviation (i.e., (post-exercise individual value – baseline mean) / baseline standard deviation), as in previous randomized controlled trials focused on cognitive outcomes ^45^. Since this variable means how many SDs the outcome has changed from baseline, we interpreted it as an effect size indicator, in which a value around 0.2 is considered a small effect size, 0.5 as a medium effect size and 0.8 as a large effect size ^46^. Discovery analyses of the exercise program on 91 neurology-related plasma proteins were adjusted for multiple comparisons using false discovery rate (FDR) based on the Benjamini-Hochberg method ^47^.

#### 2.4.2 Secondary analyses: Mediation analyses

We tested whether the effects of the exercise intervention on the brain health indicators (i.e., BDNF, hippocampal structure and function, cognitive performance and mental health) were mediated by changes in blood-circulating biomarkers, if exercise showed an effect on them. Mediation analyses were performed using R version 1.3.1073 using the Lavaan package, with a resample procedure of 5,000 bootstrap samples.

The unstandardized (B) and standardized (*β*) regression coefficients are presented for four equations: Equation 1 regressed the mediator (i.e., change in blood-circulating biomarkers) on the independent variable (group). Equation 2 regressed the dependent variables (i.e., brain health indicators) on the independent variable. Equations 3 regressed the dependent variables on both the mediator (equation 3) and the independent variable (equation 3’). The indirect effects along with its confidence intervals (CIs) were also presented and the significance was considered if the indirect effect significantly differed from zero (i.e., zero is not contained within the CIs). Finally, the percentage of total effect was computed to know how much of the total effect was explained by the mediation, as follows: (indirect effect / total effect) × 100.

#### 2.4.3 Sensitivity analyses: Intention-to-treat analyses

We tested (i.e., using ANCOVA) whether the participants that completed the baseline and post-exercise evaluations but did not attend at least 70% of the recommended 3 sessions/week, differed in the main study variables from the participants who met per-protocol criteria.

## 3. Results

The flowchart of the study is presented in **Fig. 1**. In total, 112 children with OW/OB, meeting the eligibility criteria, participated in this study. Of them, 109 were randomly allocated to an exercise group, which participated in the exercise program, or to a wait-list control group, which followed their normal life routines. A total of 22 children were excluded from analyses due to missing blood data at any time points (n=21), or poor quality control at the proteomic analyses (n=1), leaving 87 participants for the intention-to-treat analyses. Finally, 81 children with OW/OB met the per-protocol criteria (i.e., attended at least 70% of the recommended 3 sessions/week), and were included in the main analyses. Baseline sample characteristics are displayed in **Table 1**.

**Table 1.** Descriptive baseline characteristics of the *ActiveBrains* participants meeting per-protocol criteria (n=81).

|  | All |  | Exercise group |  | Control group |  |
| --- | --- | --- | --- | --- | --- | --- |
|  | N | Values | N | Values | N | Values |
| **Sex** |  |  |  |  |  |  |
| Girls (n %) | 81 | 40.7 | 42 | 35.7 | 39 | 46.2 |
| Age (years) | 81 | 10.12 (1.11) | 42 | 10.02 (1.13) | 39 | 10.23 (1.10) |
| Weight (kg) | 81 | 55.85 (11.18) | 42 | 56.61 (12.58) | 39 | 55.03 (9.54) |
| Height (cm) | 81 | 144.52 (8.39) | 42 | 143.48 (8.84) | 39 | 145.65 (7.84) |
| Body mass index (kg/m<sup>2</sup>) | 81 | 26.52 (3.56) | 42 | 27.18 (3.98) | 39 | 25.80 (2.93) |
| Peak height velocity offset (years) | 81 | -2.21 (0.97) | 42 | -2.37 (0.92) | 39 | -2.04 (1.01) |
| **Wave of participation. (%)** | 81 |  | 42 |  | 39 |  |
| First (n %) | 14 | 17.3 | 9 | 21.4 | 5 | 12.8 |
| Second (n %) | 34 | 42.0 | 17 | 40.5 | 17 | 43.6 |
| Third (n %) | 33 | 20.7 | 16 | 38.1 | 17 | 43.6 |

Values are expressed as means ± standard deviations (SD), unless otherwise indicated.

**Fig. 1.**
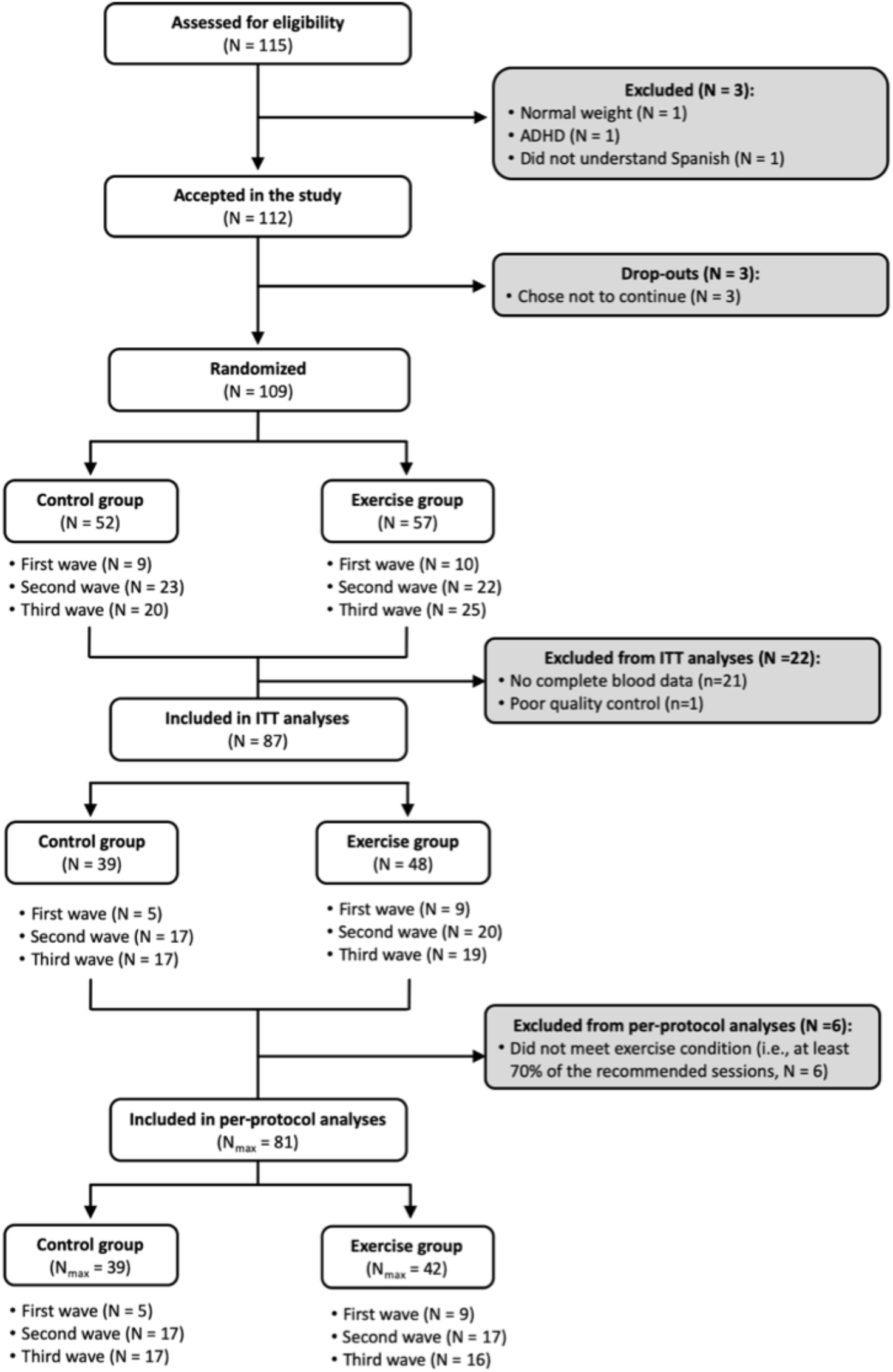
Flow chart. ITT = Intention-to-treat N_max_ = Maximum N for analyses, it changes depending on the variable, see tables X for specific sample sizes per variable.

### 3.1 Main analyses: Effects of exercise on blood-circulating biomarkers

The effects of the exercise intervention on the candidate blood-circulating biomarkers identified BDNF, BHB, CTSB, Kynurenine, FGF21, sVCAM-1 are presented in **Table 2**. Overall, no significant effects were found on any of the selected blood-circulating biomarkers (all P>0.05).

**Table 2.**
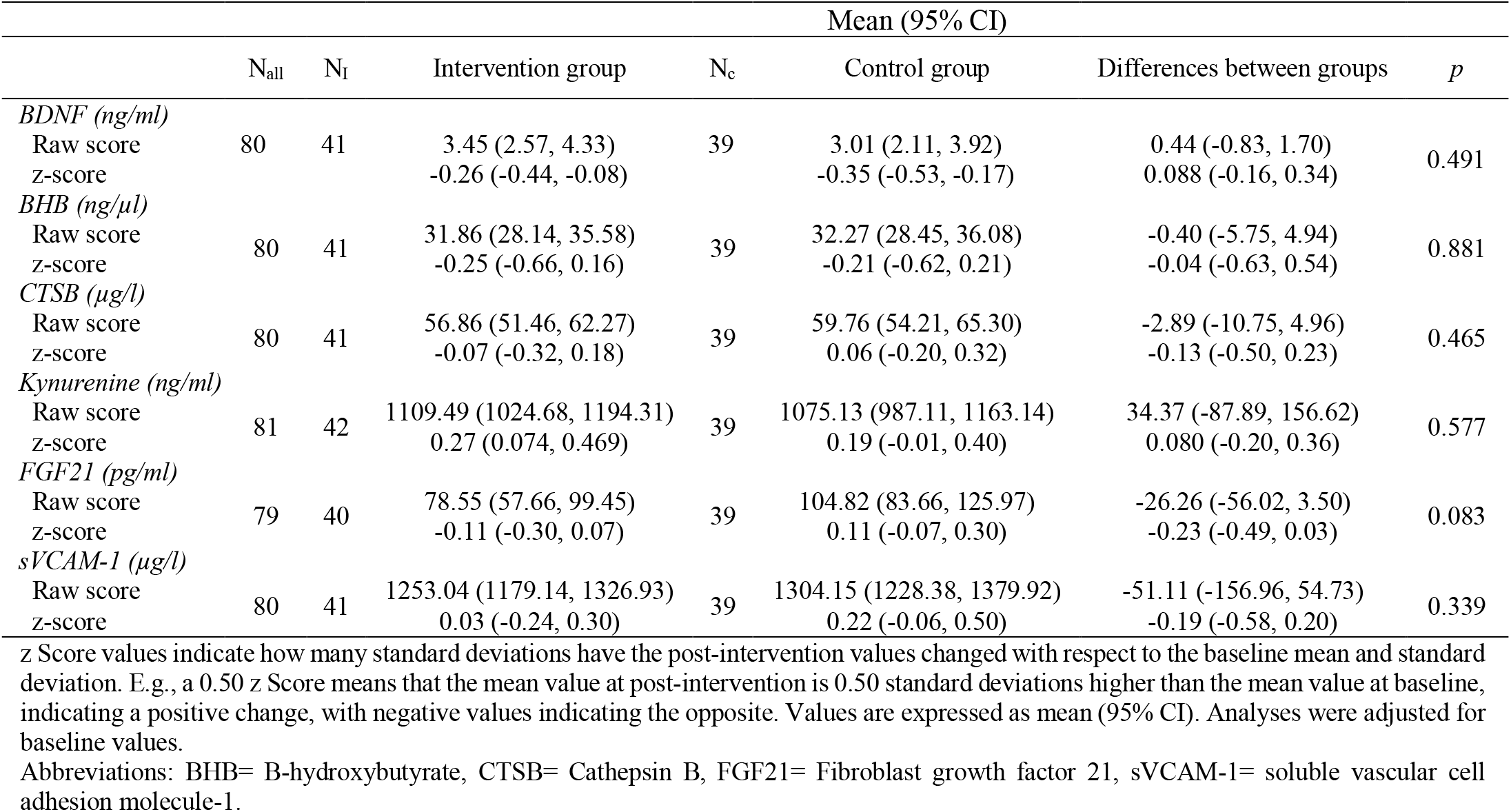
Per protocol effects of the ActiveBrains intervention on raw and z-score post-intervention candidate blood-circulating biomarkers.

|  |  |  | Mean (95% CI) |  |  |  |  |
| --- | --- | --- | --- | --- | --- | --- | --- |
|  | N<sub>all</sub> | N<sub>I</sub> | Intervention group | N<sub>c</sub> | Control group | Differences between groups | *p* |
| *BDNF (ng/ml)* |  |  |  |  |  |  |  |
| Raw score | 80 | 41 | 3.45 (2.57, 4.33) | 39 | 3.01 (2.11, 3.92) | 0.44 (-0.83, 1.70) | 0.491 |
| z-score |  |  | -0.26 (-0.44, -0.08) |  | -0.35 (-0.53, -0.17) | 0.088 (-0.16, 0.34) |  |
| *BHB (ng/μl)* |  |  |  |  |  |  |  |
| Raw score | 80 | 41 | 31.86 (28.14, 35.58) | 39 | 32.27 (28.45, 36.08) | -0.40 (-5.75, 4.94) | 0.881 |
| z-score |  |  | -0.25 (-0.66, 0.16) |  | -0.21 (-0.62, 0.21) | -0.04 (-0.63, 0.54) |  |
| *CTSB (μg/l)* |  |  |  |  |  |  |  |
| Raw score | 80 | 41 | 56.86 (51.46, 62.27) | 39 | 59.76 (54.21, 65.30) | -2.89 (-10.75, 4.96) | 0.465 |
| z-score |  |  | -0.07 (-0.32, 0.18) |  | 0.06 (-0.20, 0.32) | -0.13 (-0.50, 0.23) |  |
| *Kynurenine (ng/ml)* |  |  |  |  |  |  |  |
| Raw score | 81 | 42 | 1109.49 (1024.68, 1194.31) | 39 | 1075.13 (987.11, 1163.14) | 34.37 (-87.89, 156.62) | 0.577 |
| z-score |  |  | 0.27 (0.074, 0.469) |  | 0.19 (-0.01, 0.40) | 0.080 (-0.20, 0.36) |  |
| *FGF21 (pg/ml)* |  |  |  |  |  |  |  |
| Raw score | 79 | 40 | 78.55 (57.66, 99.45) | 39 | 104.82 (83.66, 125.97) | -26.26 (-56.02, 3.50) | 0.083 |
| z-score |  |  | -0.11 (-0.30, 0.07) |  | 0.11 (-0.07, 0.30) | -0.23 (-0.49, 0.03) |  |
| *sVCAM-1 (μg/l)* |  |  |  |  |  |  |  |
| Raw score | 80 | 41 | 1253.04 (1179.14, 1326.93) | 39 | 1304.15 (1228.38, 1379.92) | -51.11 (-156.96, 54.73) | 0.339 |
| z-score |  |  | 0.03 (-0.24, 0.30) |  | 0.22 (-0.06, 0.50) | -0.19 (-0.58, 0.20) |  |

z Score values indicate how many standard deviations have the post-intervention values changed with respect to the baseline mean and standard deviation. E.g., a 0.50 z Score means that the mean value at post-intervention is 0.50 standard deviations higher than the mean value at baseline, indicating a positive change, with negative values indicating the opposite. Values are expressed as mean (95% CI). Analyses were adjusted for baseline values.
Abbreviations: BHB= B-hydroxybutyrate, CTSB= Cathepsin B, FGF21= Fibroblast growth factor 21, sVCAM-1= soluble vascular cell adhesion molecule-1.

The effects of the exercise program on the explored 91 neurology-related proteins are presented in **Table S3A**. In addition, significant (unadjusted p-values <0.05) effects of the intervention on these neurology-related proteins are presented in **Table 3** (in detail). In addition, up- and down-regulated proteins and levels of significant is graphically represented by a volcano plot in **Fig. 2**. Overall, the exercise group reduced significantly the levels of six circulating neurology-related proteins in plasma compared to the control group, which can be interpreted as a small-to-medium effect size (ranged from -0.19 to -0.46 SDs of change, and p values from 0.001 to 0.040). Particularly, the proteins whose concentration reduced significantly after the intervention program were: (i) Carboxypeptidase A2 (CPA2), (ii) KYNU, (iii) Leukocyte-associated immunoglobulin-like receptor 2 (LAIR2), (iv) MSR1, (v) PLXNB3, and (vi) the Lysosome membrane protein 2 (SCARB2). All significant effects disappeared after correcting for multiple testing, with only the MSR1 showing a borderline corrected (FDR) P value of 0.07.

**Table 3.**
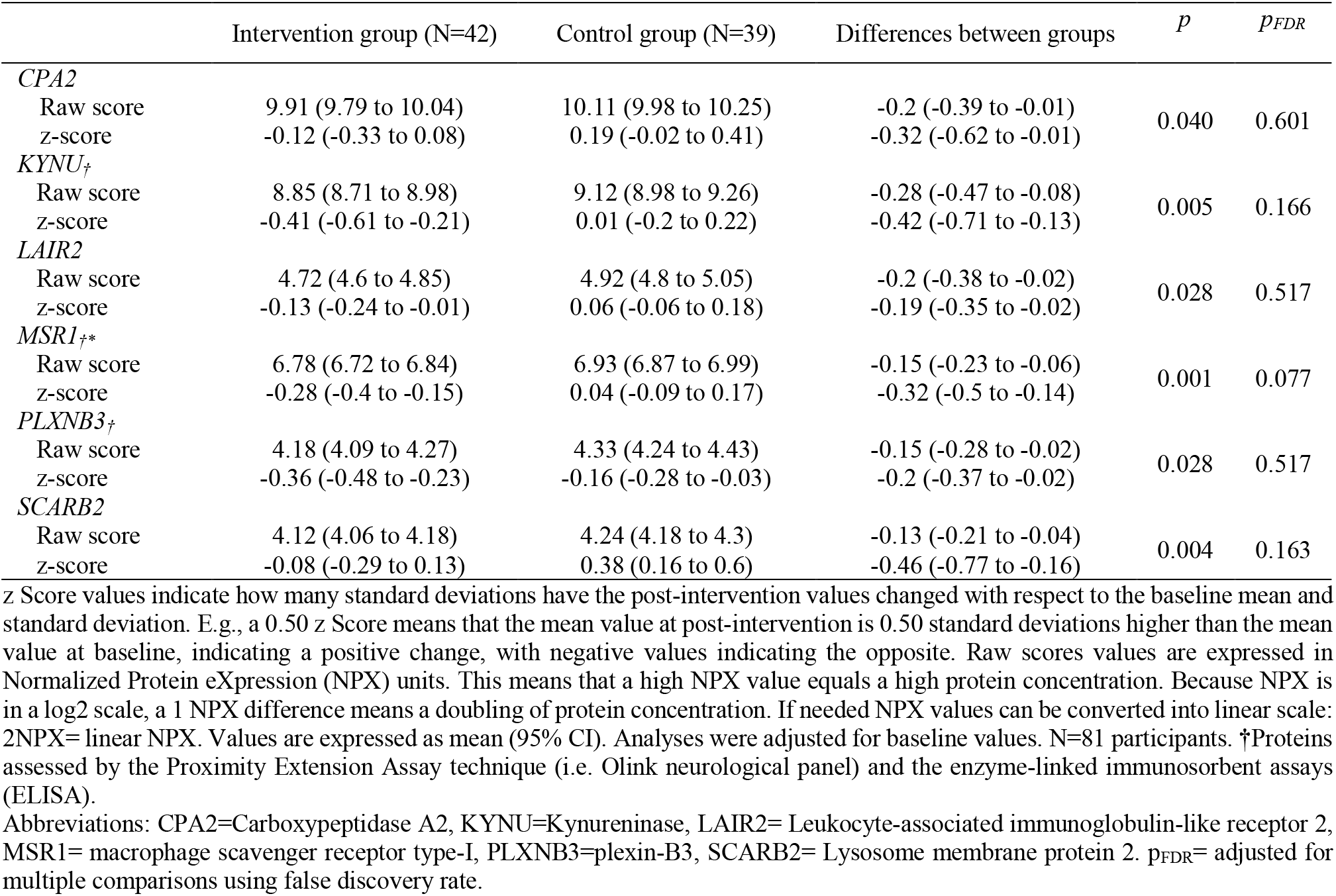
Significant per protocol effects of the ActiveBrains intervention on raw and z Score post-intervention Olink neurology-related proteins.

**Fig. 2.**
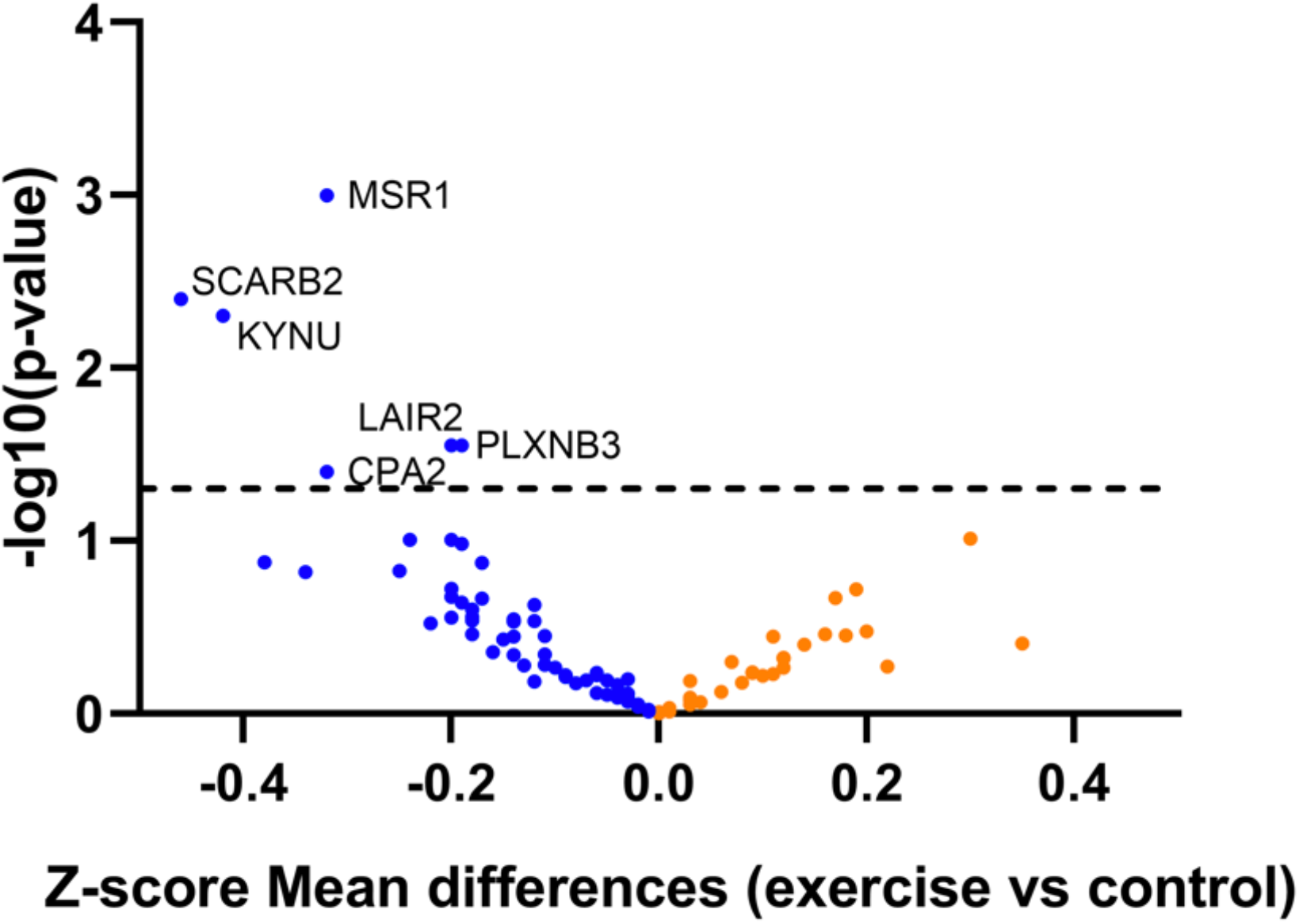
Volcano plot shows the 6 differential expressed proteins between exercise and control group in children with overweight/obesity controlling for baseline protein levels. Up-regulated proteins are in orange, while down-regulated proteins are in blue. The x-axis reflects the Normalized Protein eXpression (NPX) values mean differences (i.e., A 1 NPX value difference means a doubling of protein concentration), while the y-axis indicates statistical significance p <0.05, which is –log10 > 1.30 in the horizontal dashed line. CPA2=Carboxypeptidase A2, KYNU=Kynureninase, LAIR2= Leukocyte-associated immunoglobulin-like receptor 2, MSR1= macrophage scavenger receptor type-I, PLXNB3=plexin-B3, SCARB2= Lysosome membrane protein 2.

The validation analyses performed by ELISA assays are presented in **Table 4** (per-protocol). Briefly, three neurology-related proteins were validated with ELISA in our study (i.e., MSR1, PLXNB3, KYNU). Of them, the effect of exercise intervention was only confirmed on MSR1 (-0.33 SDs, p=0.013).

**Table 4.**
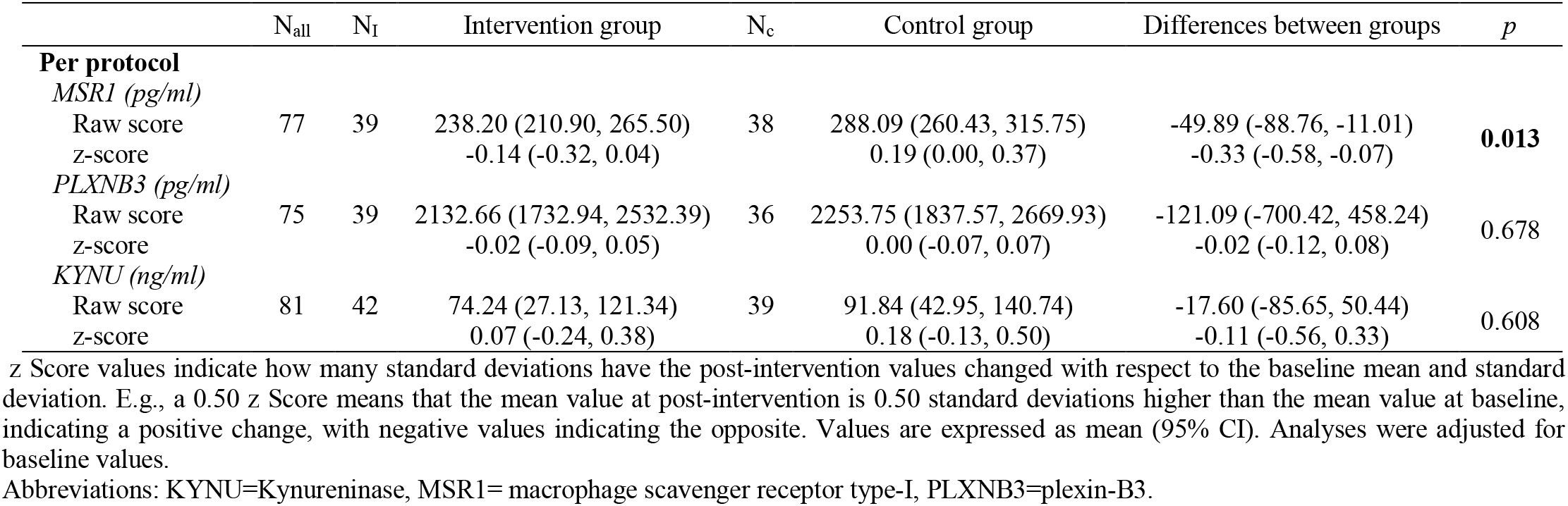
ELISA assay technique validation

### 3.2 Secondary analyses: Mediation analyses

A visualization of the mediation panel modeling approach is represented in **Fig. S1A**. Mediation analyses were performed with the per-protocol sample. The effects of the exercise intervention on the brain health outcomes, mediated by changes in neurology-related proteins (i.e., CPA2, KYNU, LAIR2, MSR1, PLXNB3, SCARB2), are presented in **Appendix 2**. Briefly, none of the neurology-related proteins mediated the chronic effects of exercise on the brain health outcomes, after correcting for FDR.

### 3.3 Sensitivity analyses: Intention-to-treat analyses

As **Table S6A** shows, the effects of exercise intervention on neurology-related proteins derived from the intention-to-treat analyses were consistent with those from the per-protocol analyses described above. The effect sizes were in overall consistent, yet slightly attenuated. This attenuation is often expected since participants who attended to few exercise sessions are often less committed to the program. These results were similar from the candidate blood-circulating biomarkers (**Table S5A**), and ELISA validation (**Table S4A**).

## 4. Discussion

The overall aim of this study was to investigate the effect of a 20-week exercise intervention in children with OW/OB on a broad set of blood-circulating biomarkers potentially related to the brain health. Overall, our exercise program did not identify any chronic effect on candidate blood-circulating biomarkers selected, i.e., BDNF, BHB, CTSB, Kynurenine, FGF21, and sVCAM-1. In the discovery analyses, we observed consistent evidence (i.e. cross-validated with two different techniques, and in per-protocol and intention-to-treat analyses) supporting that an exercise program reduces the plasma concentration of MRS1 in children with OW/OB. However, this exercise-induce change in MRS1 did not seem to mediate the chronic effect of exercise on BDNF, hippocampus structure and function, cognition, and mental health in children with OW/OB.

This study did not confirm that exercise affects blood-circulating biomarkers previously identified as potential molecular mechanisms through which exercise might improve brain health in children. There are several possible explanations. First, most previous studies investigated these molecular pathways in rodents ^23^. Second, the recent controversial studies developed in humans were performed mainly in healthy adults ^22,48–50^ or unhealthy populations (e.g., people with type-II diabetes or major depression) ^31,51–53^, which preclude to compare our findings in pediatric population. Third, the use of different assessment methodologies (e.g., biological medium, centrifugation strategy, temperature, and choice of bioassay) may introduce bias and complicate comparisons between studies. Consequently, future studies should standardize the measurement methodology of these biomarkers to improve comparability between studies. Lastly, most of the available evidence in humans about the effect of exercise on circulating blood levels of the above mentioned biomarkers has been studied in acute exercise, while we investigated the effect of chronic exercise. For instance, it is well known the acute effect of exercise on circulating blood FGF21 levels, although the effect of chronic exercise has not been so widely studied ^54^. Furthermore, it is possible that we did not observe any effect since the most of the selected biomarkers are short-life molecules, and consequently, they returned to basal levels within minutes or hours after the stimuli.

The most robust and consistent finding of the present study is that a 20-week exercise program reduced the protein levels of MSR1 in the plasma of children with OW/OB. MSR1 has not been related to exercise before. The MSR1 is a membrane glycoprotein, expressed in macrophages, which is involved in pathological processes such as atherosclerosis, non-alcoholic fatty liver disease, and neurological diseases (e.g., Alzheimer’s disease and multiple sclerosis) ^55–58^. Concerning atherosclerosis, oxidized lipoproteins (oxLDL) are one of the main factors that contribute to the initiation of atherosclerosis ^59,60^, while scavenger receptors such as MSR1 lead macrophages accumulate modified lipoproteins (e.g., oxLDL) and contribute to the formation of atherosclerotic plaque ^55^. In this regard, the late stage of atherosclerosis is characterized by an increased monocyte and macrophage activity, and systemic inflammation ^55^. Interestingly, re-analysis of subjects after 6 months of treatment, showed a significant decrease in the MSR1 expression in all patients who received atorvastatin. The results of presented studies demonstrate that both investigated receptors are involved in the development and/or progression of Acute Coronary Syndromes ^55^. Thus, *MSR1* up-regulation in circulating monocytes of patients with acute coronary syndrome might be associated with the late stage of atherosclerosis and inflammation. Concerning non-alcoholic fatty liver disease, Govaere et al. recently found that higher levels of MSR1 were associated with greater accumulation in Kupffer cells (the endogenous hepatic macrophages) resulting in liver inflammation, and in turn, promoting the progression of non-alcoholic fatty liver disease in humans and in animal models ^58^. Taken together, we showed that a 20-week exercise intervention is effective to reduce the circulating plasma levels of MSR1 in children with OW/OB, which might improve some cardiovascular risk factors (e.g., inflammatory profile or lipid levels) associated with the development of subclinical atherosclerosis in youth ^61,62^, and other neurological diseases (e.g., Alzheimer’s disease and multiple sclerosis) later in life.

In the context of neurological disorders, Frenkel et al. reported that amyloid-β (Aβ) accumulation in the brain was positively associated with MSR1 deficiency in mice. While, Aβ clearance was increased after pharmacological up-regulation of MSR1 in vitro, which could attenuate Alzheimer’s disease progression^56^. On the other hand, in multiple sclerosis, the *MSR1* gene was up-regulated in chronic active lesions (post-mortem human brain) compared to control tissues and could contribute to demyelination ^63^. Besides, central nervous system demyelination was reduced in Msr1-/- mice (i.e., deficient in MSR1) compared to wild-type ^56^. In relation to “exercise”, one study reported that the *MSR1* gene was down-regulated in the blood of older adults with prostate cancer that did not practice sports compared to “active” patients ^64^. In summary, the heterogeneity among studies (e.g., humans or rodents; different tissues analyzed; reported MSR1 protein or *MSR1* gene) and the lack of studies about exercise and MSR1 make difficult the interpretation of the direction of changes in MSR1 (i.e., chronic exercise reduced the MSR1 expression in plasma) in our study. In addition, MSR1 changes did not mediate the chronic exercise effects on brain health indicators. However, chronic exercise decreased MSR1 protein in plasma of children with OW/OB performing two different techniques (i.e., Olink and ELISA), reporting similar effect sizes. Thus, future studies should study the effects of exercise (chronic or acute) on MSR1 protein expression in different tissues and combining humans (e.g., different age ranges and clinical conditions) and animal models to reveal the molecular mechanisms underlying the effect of exercise on brain health indicators. In this regard, MSR1 protein may be considered a novel candidate for further studies.

Blood-circulating biomarkers, in which our exercise program had small effects, did not mediate the effect of exercise on brain health indicators during childhood. Noteworthy is that our exercise intervention has almost no effects on most of the brain health outcomes included in this study ^39^. Therefore, the unmediated role of the explored biomarkers may be partially explained by the small effect of our exercise program on the brain health of children with OW/OB. Additionally, other blood-circulating biomarkers, not included in this study, could be mediating the effect of exercise on brain health during childhood. For instance, irisin (i.e., a muscle-derived factor, secreted from muscle after shedding of the extracellular portion of the type I membrane protein FNDC5) seems to be increased after exercise in the hippocampus of mice ^20^. However, the measurement of irisin with ELISA methods remains challenging and controversial in humans ^65,66^. In fact, we decided not to measure Irisin in our study due to the lack of specificity in the currently available ELISA methods claimed by numerous authors ^67^. Moreover, more studies are need to delineate the neurobiological, but also psychosocial, and behavioral mechanisms for the effect of exercise on brain health in childhood ^68^.

A strength of the present study is its design itself, which guarantee that the only difference between two treatments (exercise vs. control) is their exposure to the treatment of interest. Moreover, much of the previous evidence on the role of exercise in blood-circulating biomarkers is based on acute-effect studies. Our study provides evidence on the chronic effects of exercise on these biomarkers which is novel and has implications for people practicing exercise on a regular basis. Another strength of this study is the inclusion of a broad set of novel blood-circulating biomarkers, including a candidate-based approach, as well as a discovery approach on a large number of proteins involved in neurological processes. Our study has also a few limitations that need to be considered. Although precautions were taken to reduce the risk of bias in the evaluations (e.g., randomization after baseline assessment, physical trainers not involved in the evaluations), some of the project staff involved in the post-exercise evaluations (e.g., cognitive and mental health assessment) were not blinded to the group allocation which could add some bias to the post-intervention measurements. However, this does not influence the main findings of this study, since all blood-circulating biomarkers were analyzed by personnel blinded to the group allocation, and so apply to the MRI outcomes. Our study was conducted in children with OW/OB, and it is unknown the extent to which our findings apply to other populations.

## 5. Conclusion

Our study does not support a chronic effect of exercise on studied candidate biomarkers of brain health in children with OW/OV. However, our discover analysis identified a potential reduction effect of exercise on levels of blood-circulating MRS1, which was validated using ELISA methods. Lastly, exercise-induced changes in MRS1 or any other biomarker studied did not mediated changes in brain health indicators. Future studies will confirm or contrast the effect of exercise on MRS1 and its implications on health outcomes.

## Data Availability

All data produced in the present study are available upon reasonable request to the authors

## Acknowledgments

This work study was mainly supported by the Spanish Ministry of Economy and Competitiveness (DEP2017-91544-EXP) and the Alicia Koplowitz Foundation. This study was also supported by grants from the Spanish Ministry of Economy and Competitiveness (DEP2013-47540, DEP2016-79512-R), European Regional Development Fund (ERDF)”, and by the European Commission (No 667302). Additional funding was obtained from the Andalusian Operational Programme supported with ERDF (FEDER in Spanish, B-CTS-355-UGR18, B-CTS-500-UGR18 and A-CTS-614-UGR20). This study was partially funded by the University of Granada, Plan Propio de Investigación 2016, Excellence actions: Units of Excellence; Unit of Excellence on Exercise and Health (UCEES) and by the Junta de Andalucía, Consejería de Conocimiento, Investigación y Universidades and ERDF (SOMM17/6107/UGR). In addition, this study was further supported by the SAMID III network, RETICS, funded by the PN I+D+I 2017-2021 (Spain). MRA was funded by the Ramón Areces Foundation, DEP2017-91544-EXP, the Alicia Koplowitz Foundation (ALICIAK-2018). AMG with FPU16/ 03653. IEC is supported by the Spanish Ministry of Science and Innovation (RYC2019-027287-I). PSU is supported by a grant from ANID/BECAS Chile/72180543 and through a Margarita Salas grant from the Spanish Ministry Universities. SA is supported by the Spanish Ministry of Economy, Industry and Competitiveness (MINECO) and European Regional Development Fund (FEDER): grants RYC-2016-21199 and SAF2017-87526-R, and the Junta de Andalucía (PAIDI P20_00158). APF is supported by the Spanish Ministry of Education, Culture and Sport (FPU 16/02760). The authors report no biomedical financial interests or potential conflicts of interest.

## Authors’ contributions

MR-A participated in the study design/conception, data acquisition, data analysis, interpretation, manuscript preparing and revision, and the final approval of the manuscript; FBO participated in the design/conception, data acquisition, data analysis, interpretation, manuscript preparing and revision, and the final approval of the manuscript; IE-C participated in the design/conception, data acquisition, data analysis, interpretation, manuscript preparing and revision, and the final approval of the manuscript; AP-F participated in the data acquisition, data analysis, interpretation, manuscript preparing and revision, and the final approval of the manuscript; AM-G participated in the data acquisition, interpretation, manuscript preparing and revision, and the final approval of the manuscript; PS-U participated in the data acquisition, data analysis, interpretation, manuscript preparing and revision, and the final approval of the manuscript; SA participated in the data analysis, interpretation, manuscript preparing and revision, and the final approval of the manuscript; CMA participated in the interpretation, manuscript preparing and revision, and the final approval of the manuscript; AC participated in the interpretation, manuscript preparing and revision, and the final approval of the manuscript. All authors agreed to be accountable for all aspects of work, contributed to the manuscript writing, and read and approved the final version of this manuscript.

