## Appendix 1 for "The effects of a 20-week exercise program on blood-circulating biomarkers related to brain health in children with overweight or obesity: The ActiveBrains project"

**Appendix A**

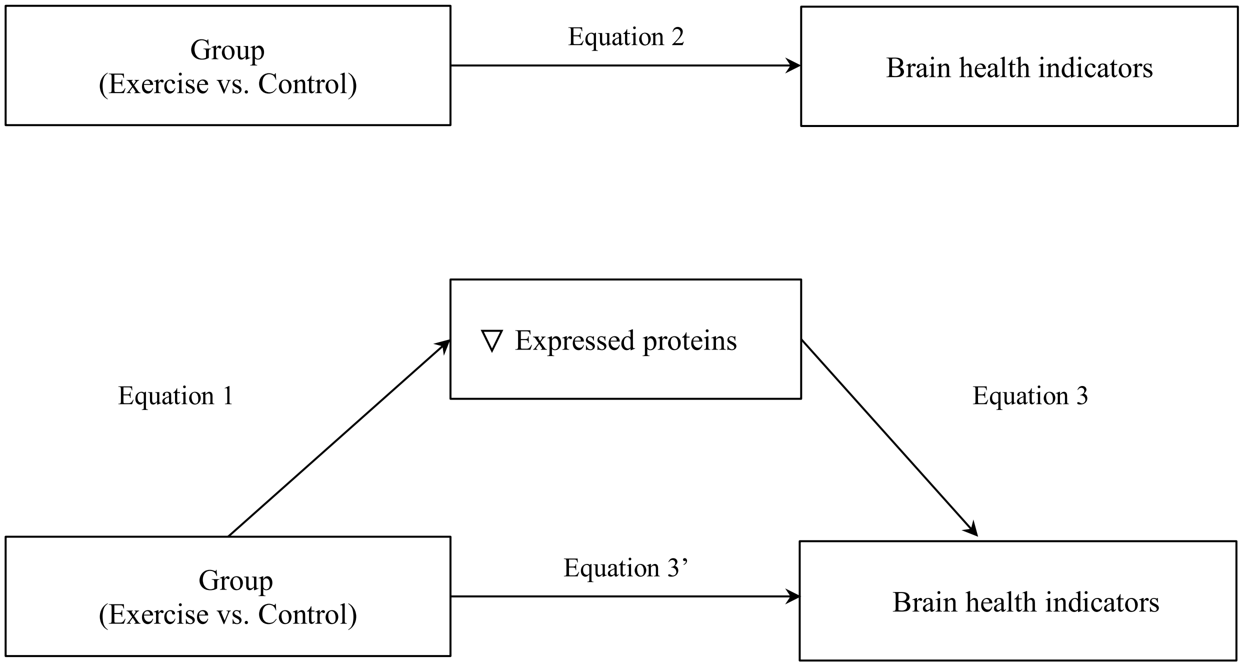


**Fig. S1A.** Visualization of the mediation panel modeling approach. Expressed proteins= Kynureninase, Leukocyte-associated immunoglobulin-like receptor 2, macrophage scavenger receptor type-I, plexin-B3, and Lysosome membrane protein 2. Brain health indicators= brain-derived neurotrophic factor, mental health indicators, cognitive performance indicators, hippocampal volume and function.

**Table S1A.** List of proteins included in the Olink Target Neurology panel.

ADP-ribosyl cyclase/cyclic ADP-ribose hydrolase 1 (CD38)
Alpha-2-macroglobulin receptor-associated protein (Alpha-2-MRAP)
BDNF/NT-3 growth factors receptor (NTRK2)
Beta-nerve growth factor (Beta-NGF)
Bone morphogenetic protein 4 (BMP-4)
Brevican core protein (BCAN)
Brorin (VWC2)
C-type lectin domain family 1 member B (CLEC1B)
C-type lectin domain family 10 member A (CLEC10A)
Cadherin-3 (CDH3)
Cadherin-6 (CDH6)
Carboxypeptidase A2 (CPA2)
Carboxypeptidase M (CPM)
Cathepsin S (CTSS)
Cell adhesion molecule 3 (CADM3)
Cell surface glycoprotein CD200 receptor 1 (CD200R1)
CMRF35-like molecule 1 (CLM-1)
CMRF35-like molecule 6 (CLM-6)
Contactin-5 (CNTN5)
Cytotoxic and regulatory T-cell molecule (CRTAM)
Dickkopf-related protein 4 (Dkk-4)
Dipeptidyl peptidase 1 (CTSC)
Disintegrin and metalloproteinase domain-containing protein 22 (ADAM 22)
Disintegrin and metalloproteinase domain-containing protein 23 (ADAM 23)
Draxin (DRAXIN)
Ephrin type-B receptor 6 (EPHB6)
Ephrin-A4 (EFNA4)
Epithelial discoidin domain-containing receptor 1 (DDR1)
Ezrin (EZR)
Fc receptor-like protein 2 (FcRL2)
Galectin-8 (gal-8)
GDNF family receptor alpha-1 (GFR-alpha-1)
GDNF family receptor alpha-3 (GDNFR-alpha-3)
Glial cell line-derived neurotrophic factor (GDNF)
Glypican-5 (GPC5)
Granulocyte Colony-Stimulating Factor (G-CSF)
Granulocyte-macrophage colony-stimulating factor receptor subunit alpha (GM-CSF-R-alpha)
Granzyme A (GZMA)
Growth/differentiation factor 8 (GDF-8)
Hydroxyacylglutathione hydrolase, mitochondrial (HAGH)
Interleukin-12 subunit beta, Interleukin-12 subunit alpha (IL-12B, IL-12A)
Interleukin-5 receptor subunit alpha (IL-5R-alpha)
Junctional adhesion molecule B (JAM-B)
Kynureninase (KYNU)
Latexin (LXN)
Layilin (LAYN)
Leucine-rich repeat transmembrane protein FLRT2 (FLRT2)
Leukocyte-associated immunoglobulin-like receptor 2 (LAIR-2)
Linker for activation of T-cells family member 1 (LAT)
Lysosome membrane protein 2 (SCARB2)
Macrophage scavenger receptor types I and II (MSR1)
MAM domain-containing glycosylphosphatidylinositol anchor protein 1 (MDGA1)
Matrilin-3 (MATN3)
Mesencephalic astrocyte-derived neurotrophic factor (MANF)
Microtubule-associated protein tau (MAPT)
N-acylethanolamine-hydrolyzing acid amidase (NAAA)
Neprilysin (NEP)
Netrin receptor UNC5C (UNC5C)

| |
|---|
| Neuroblastoma suppressor of tumorigenicity 1 (NBL1) |
| Neurocan core protein (NCAN) |
| Neuronal cell adhesion molecule (Nr-CAM) |
| Neuropilin-2 (NRP2) |
| Neutral ceramidase (N-CDase) |
| Nicotinamide/nicotinic acid mononucleotide adenylyltransferase 1 (NMNAT1) |
| NKG2D ligand 2 (N2DL-2) |
| NT-3 growth factor receptor (NTRK3) |
| OX-2 membrane glycoprotein (CD200) |
| Platelet-derived growth factor receptor alpha (PDGF-R-alpha) |
| Plexin-B1 (PLXNB1) |
| Plexin-B3 (PLXNB3) |
| Poliovirus receptor (PVR) |
| Protogenin (PRTG) |
| R-spondin-1 (RSPO1) |
| Repulsive guidance molecule A (RGMA) |
| RGM domain family member B (RGMB) |
| Roundabout homolog 2 (ROBO2) |
| Scavenger receptor class A member 5 (SCARA5) |
| Scavenger receptor class F member 2 (SCARF2) |
| Secreted frizzled-related protein 3 (sFRP-3) |
| Serine/threonine-protein kinase receptor R3 (SKR3) |
| Sialic acid-binding Ig-like lectin 9 (Siglec-9) |
| Sialoadhesin (SIGLEC1) |
| SPARC-related modular calcium-binding protein 2 (SMOC2) |
| Sphingomyelin phosphodiesterase (SMPD1) |
| Tenascin-R (TN-R) |
| Testican-1 (SPOCK1) |
| Thy-1 membrane glycoprotein (THY 1) |
| Transmembrane protease serine 5 (TMPRSS5) |
| Tumor necrosis factor receptor superfamily member 12A (TNFRSF12A) |
| Tumor necrosis factor receptor superfamily member 21 (TNFRSF21) |
| Tumor necrosis factor receptor superfamily member 27 (EDA2R) |
| WAP, Kazal, immunoglobulin, Kunitz and NTR domain-containing protein 1 (WFIKKN1) |

**Table S2A.** Baseline partial correlations adjusted for sex and peak height velocity between BDNF and the 6 differential expressed proteins between exercise and control group in children with overweight/obesity.

| | | SCARB2 | MSR1 | KYNU | LAIR | CPA2 | PLXNB3 |
|---|---|---|---|---|---|---|---|
| BDNF | r | -0.043 | 0.001 | 0.563 | 0.018 | 0.146 | 0.757 |
| | *p* | 0.705 | 0.990 | <0.001 | 0.877 | 0.202 | <0.001 |

Abbreviations: BDNF= brain-derived neurotrophic factor, CPA2=Carboxypeptidase A2, KYNU=Kynureninase, LAIR2= Leukocyte-associated immunoglobulin-like receptor 2, MSR1= macrophage scavenger receptor type-I, PLXNB3=plexin-B3, SCARB2= Lysosome membrane protein 2.

**Table S3A.** Per protocol effects of the ActiveBrains intervention on raw and z Score post-intervention neurology-related proteins (n=81)

| | Intervention group | Control group | Differences between groups | *p* | *p*$_{FDR}$ |
|---|---|---|---|---|---|
| ADAM22 | | | | | |
| Raw score | 4.99 (4.91 to 5.07) | 4.98 (4.9 to 5.06) | 0.01 (-0.1 to 0.12) | 0.887 | 0.972 |
| z-score | 0.09 (-0.19 to 0.36) | 0.06 (-0.23 to 0.34) | 0.03 (-0.37 to 0.42) | | |
| ADAM23 | | | | | |
| Raw score | 4.95 (4.88 to 5.02) | 4.91 (4.84 to 4.99) | 0.04 (-0.07 to 0.14) | 0.475 | 0.891 |
| z-score | -0.07 (-0.3 to 0.15) | -0.19 (-0.43 to 0.04) | 0.12 (-0.21 to 0.45) | | |
| Alpha2MRAP | | | | | |
| Raw score | 8.02 (7.84 to 8.19) | 8.04 (7.86 to 8.22) | -0.03 (-0.28 to 0.23) | 0.834 | 0.967 |
| z-score | -0.2 (-0.38 to -0.02) | -0.17 (-0.36 to 0.02) | -0.03 (-0.29 to 0.24) | | |
| BCAN | | | | | |
| Raw score | 5.04 (4.98 to 5.1) | 5.08 (5.02 to 5.15) | -0.04 (-0.13 to 0.05) | 0.359 | 0.884 |
| z-score | -0.32 (-0.52 to -0.12) | -0.18 (-0.39 to 0.03) | -0.14 (-0.43 to 0.16) | | |
| BetaNGF | | | | | |
| Raw score | 1.28 (1.24 to 1.32) | 1.29 (1.25 to 1.33) | -0.01 (-0.06 to 0.05) | 0.776 | 0.962 |
| z-score | -0.21 (-0.4 to -0.02) | -0.17 (-0.37 to 0.02) | -0.04 (-0.31 to 0.23) | | |
| BMP4 | | | | | |
| Raw score | 4.45 (4.37 to 4.53) | 4.45 (4.37 to 4.53) | 0 (-0.11 to 0.11) | 0.972 | 0.995 |
| z-score | 0.02 (-0.24 to 0.29) | 0.03 (-0.24 to 0.3) | -0.01 (-0.39 to 0.38) | | |
| CADM3 | | | | | |
| Raw score | 4.82 (4.74 to 4.9) | 4.83 (4.74 to 4.91) | -0.01 (-0.13 to 0.11) | 0.885 | 0.972 |
| z-score | -0.15 (-0.34 to 0.04) | -0.13 (-0.33 to 0.06) | -0.02 (-0.29 to 0.25) | | |
| CD200 | | | | | |
| Raw score | 7.06 (7 to 7.12) | 7.06 (6.99 to 7.12) | 0 (-0.09 to 0.09) | 0.995 | 0.995 |
| z-score | -0.12 (-0.36 to 0.13) | -0.12 (-0.37 to 0.13) | 0 (-0.35 to 0.35) | | |
| CD200R1 | | | | | |
| Raw score | 5.13 (5.08 to 5.18) | 5.17 (5.12 to 5.22) | -0.04 (-0.1 to 0.03) | 0.291 | 0.884 |
| z-score | -0.18 (-0.33 to -0.02) | -0.06 (-0.22 to 0.1) | -0.12 (-0.34 to 0.1) | | |
| CD38 | | | | | |
| Raw score | 5.95 (5.88 to 6.01) | 6 (5.93 to 6.07) | -0.05 (-0.15 to 0.05) | 0.301 | 0.884 |
| z-score | -0.32 (-0.62 to -0.02) | -0.09 (-0.4 to 0.21) | -0.22 (-0.65 to 0.2) | | |
| CDH3 | | | | | |
| Raw score | 8.05 (7.98 to 8.12) | 8.12 (8.04 to 8.19) | -0.07 (-0.17 to 0.03) | 0.190 | 0.884 |
| z-score | -0.12 (-0.33 to 0.09) | 0.09 (-0.14 to 0.31) | -0.2 (-0.51 to 0.1) | | |
| CDH6 | | | | | |
| Raw score | 5.4 (5.36 to 5.45) | 5.38 (5.34 to 5.43) | 0.02 (-0.05 to 0.08) | 0.605 | 0.891 |
| z-score | -0.09 (-0.36 to 0.19) | -0.19 (-0.47 to 0.1) | 0.1 (-0.29 to 0.5) | | |
| CLEC10A | | | | | |
| Raw score | 5.61 (5.53 to 5.7) | 5.69 (5.6 to 5.78) | -0.08 (-0.2 to 0.05) | 0.212 | 0.884 |
| z-score | -0.13 (-0.36 to 0.09) | 0.07 (-0.16 to 0.3) | -0.2 (-0.53 to 0.12) | | |
| CLEC1B | | | | | |
| Raw score | 9.8 (9.54 to 10.07) | 9.9 (9.63 to 10.18) | -0.1 (-0.48 to 0.28) | 0.600 | 0.891 |
| z-score | -0.32 (-0.55 to -0.09) | -0.23 (-0.47 to 0.01) | -0.09 (-0.42 to 0.24) | | |
| CLM1 | | | | | |
| Raw score | 6.3 (6.21 to 6.38) | 6.33 (6.24 to 6.42) | -0.03 (-0.15 to 0.1) | 0.642 | 0.891 |
| z-score | -0.08 (-0.22 to 0.07) | -0.03 (-0.18 to 0.12) | -0.05 (-0.26 to 0.16) | | |
| CLM6 | | | | | |
| Raw score | 5.79 (5.75 to 5.83) | 5.83 (5.79 to 5.87) | -0.04 (-0.1 to 0.01) | 0.134 | 0.884 |
| z-score | -0.2 (-0.36 to -0.04) | -0.03 (-0.2 to 0.14) | -0.17 (-0.41 to 0.06) | | |
| CNTN5 | | | | | |
| Raw score | 6.2 (6.11 to 6.28) | 6.16 (6.07 to 6.25) | 0.03 (-0.09 to 0.16) | 0.583 | 0.891 |
| z-score | -0.24 (-0.46 to -0.02) | -0.33 (-0.56 to -0.1) | 0.09 (-0.23 to 0.41) | | |
| **CPA2** | | | | | |
| Raw score | 9.91 (9.79 to 10.04) | 10.11 (9.98 to 10.25) | -0.2 (-0.39 to -0.01) | **0.040** | 0.601 |
| z-score | -0.12 (-0.33 to 0.08) | 0.19 (-0.02 to 0.41) | -0.32 (-0.62 to -0.01) | | |
| CPM | | | | | |
| Raw score | 7.06 (7.01 to 7.1) | 7.05 (7 to 7.09) | 0.01 (-0.05 to 0.07) | 0.807 | 0.966 |
| z-score | -0.01 (-0.19 to 0.17) | -0.04 (-0.23 to 0.14) | 0.03 (-0.23 to 0.29) | | |
| CRTAM | | | | | |
| Raw score | 5.92 (5.85 to 5.99) | 5.95 (5.88 to 6.02) | -0.03 (-0.13 to 0.07) | 0.584 | 0.891 |
| z-score | -0.21 (-0.35 to -0.07) | -0.15 (-0.3 to -0.01) | -0.06 (-0.26 to 0.14) | | |
| CTSC | | | | | |
| Raw score | 2.95 (2.85 to 3.04) | 2.86 (2.76 to 2.96) | 0.09 (-0.05 to 0.22) | 0.214 | 0.884 |
| z-score | 0 (-0.19 to 0.19) | -0.17 (-0.36 to 0.02) | 0.17 (-0.1 to 0.44) | | |

**Table S3A.** (Continued)

| | | | | | |
|---|---|---|---|---|---|
| CTSS | | | | | |
| Raw score | 5.67 (5.62 to 5.72) | 5.61 (5.56 to 5.66) | 0.06 (-0.01 to 0.13) | 0.097 | 0.884 |
| z-score | 0.2 (-0.05 to 0.44) | -0.1 (-0.36 to 0.15) | 0.3 (-0.06 to 0.65) | | |
| DDR1 | | | | | |
| Raw score | 7.74 (7.69 to 7.78) | 7.75 (7.71 to 7.8) | -0.02 (-0.08 to 0.05) | 0.612 | 0.891 |
| z-score | -0.25 (-0.49 to -0.01) | -0.16 (-0.41 to 0.08) | -0.09 (-0.43 to 0.26) | | |
| Dkk4 | | | | | |
| Raw score | 2.36 (2.24 to 2.49) | 2.5 (2.37 to 2.63) | -0.14 (-0.32 to 0.04) | 0.133 | 0.884 |
| z-score | -0.1 (-0.44 to 0.25) | 0.29 (-0.07 to 0.64) | -0.38 (-0.89 to 0.12) | | |
| DRAXIN | | | | | |
| Raw score | 4.24 (4.15 to 4.34) | 4.25 (4.15 to 4.34) | 0 (-0.13 to 0.13) | 0.982 | 0.995 |
| z-score | -0.09 (-0.28 to 0.11) | -0.09 (-0.29 to 0.12) | 0 (-0.29 to 0.28) | | |
| EDA2R | | | | | |
| Raw score | 3.74 (3.67 to 3.82) | 3.77 (3.69 to 3.84) | -0.03 (-0.13 to 0.08) | 0.643 | 0.891 |
| z-score | 0.18 (-0.04 to 0.4) | 0.26 (0.03 to 0.49) | -0.07 (-0.39 to 0.24) | | |
| EFNA4 | | | | | |
| Raw score | 3.52 (3.47 to 3.56) | 3.57 (3.52 to 3.62) | -0.05 (-0.12 to 0.02) | 0.150 | 0.884 |
| z-score | -0.16 (-0.4 to 0.08) | 0.09 (-0.15 to 0.34) | -0.25 (-0.6 to 0.09) | | |
| EPHB6 | | | | | |
| Raw score | 4.92 (4.86 to 4.97) | 4.93 (4.87 to 4.98) | -0.01 (-0.09 to 0.07) | 0.817 | 0.966 |
| z-score | -0.26 (-0.46 to -0.06) | -0.22 (-0.43 to -0.02) | -0.03 (-0.32 to 0.26) | | |
| EZR | | | | | |
| Raw score | 5.11 (5.04 to 5.18) | 5.16 (5.09 to 5.24) | -0.06 (-0.16 to 0.05) | 0.279 | 0.884 |
| z-score | 0.05 (-0.2 to 0.3) | 0.25 (-0.01 to 0.51) | -0.2 (-0.57 to 0.17) | | |
| FcRL2 | | | | | |
| Raw score | 5.35 (5.28 to 5.41) | 5.42 (5.36 to 5.49) | -0.08 (-0.17 to 0.02) | 0.104 | 0.884 |
| z-score | -0.2 (-0.36 to -0.05) | -0.02 (-0.18 to 0.14) | -0.19 (-0.41 to 0.04) | | |
| FLRT2 | | | | | |
| Raw score | 3.23 (3.16 to 3.3) | 3.25 (3.18 to 3.32) | -0.02 (-0.12 to 0.07) | 0.655 | 0.891 |
| z-score | -0.09 (-0.45 to 0.26) | 0.02 (-0.35 to 0.4) | -0.12 (-0.64 to 0.4) | | |
| gal8 | | | | | |
| Raw score | 5.7 (5.58 to 5.82) | 5.74 (5.61 to 5.86) | -0.04 (-0.21 to 0.14) | 0.682 | 0.900 |
| z-score | -0.27 (-0.4 to -0.14) | -0.23 (-0.37 to -0.1) | -0.04 (-0.22 to 0.15) | | |
| GCP5 | | | | | |
| Raw score | 5.32 (5.17 to 5.47) | 5.24 (5.09 to 5.4) | 0.08 (-0.14 to 0.29) | 0.475 | 0.891 |
| z-score | -0.18 (-0.41 to 0.04) | -0.3 (-0.54 to -0.07) | 0.12 (-0.21 to 0.44) | | |
| GCSF | | | | | |
| Raw score | 3.62 (3.46 to 3.78) | 3.73 (3.56 to 3.89) | -0.11 (-0.34 to 0.12) | 0.347 | 0.884 |
| z-score | -0.27 (-0.53 to -0.01) | -0.09 (-0.36 to 0.18) | -0.18 (-0.55 to 0.2) | | |
| GDF8 | | | | | |
| Raw score | 4.26 (4.15 to 4.36) | 4.22 (4.11 to 4.32) | 0.04 (-0.11 to 0.19) | 0.591 | 0.891 |
| z-score | 0.12 (-0.16 to 0.39) | 0.01 (-0.28 to 0.29) | 0.11 (-0.29 to 0.5) | | |
| GDNF | | | | | |
| Raw score | 1.36 (1.27 to 1.45) | 1.36 (1.27 to 1.46) | 0 (-0.13 to 0.13) | 0.995 | 0.995 |
| z-score | 0.02 (-0.35 to 0.39) | 0.02 (-0.37 to 0.4) | 0 (-0.53 to 0.53) | | |
| GDNFRalpha3 | | | | | |
| Raw score | 5.54 (5.48 to 5.61) | 5.54 (5.47 to 5.6) | 0.01 (-0.08 to 0.1) | 0.840 | 0.967 |
| z-score | -0.06 (-0.29 to 0.17) | -0.09 (-0.33 to 0.15) | 0.03 (-0.3 to 0.37) | | |
| GFRalpha1 | | | | | |
| Raw score | 7.56 (7.5 to 7.62) | 7.56 (7.5 to 7.62) | 0 (-0.09 to 0.08) | 0.951 | 0.995 |
| z-score | -0.12 (-0.35 to 0.12) | -0.11 (-0.35 to 0.14) | -0.01 (-0.35 to 0.33) | | |
| GMCSFRalpha | | | | | |
| Raw score | 6.1 (6.04 to 6.16) | 6.12 (6.06 to 6.18) | -0.02 (-0.1 to 0.06) | 0.632 | 0.891 |
| z-score | -0.08 (-0.17 to 0) | -0.05 (-0.14 to 0.04) | -0.03 (-0.15 to 0.09) | | |
| GZMA | | | | | |
| Raw score | 6.35 (6.23 to 6.47) | 6.45 (6.33 to 6.57) | -0.1 (-0.27 to 0.07) | 0.251 | 0.884 |
| z-score | -0.1 (-0.32 to 0.11) | 0.07 (-0.15 to 0.3) | -0.18 (-0.49 to 0.13) | | |
| HAGH | | | | | |
| Raw score | 6.29 (5.96 to 6.61) | 6.14 (5.8 to 6.48) | 0.15 (-0.32 to 0.62) | 0.536 | 0.891 |
| z-score | 1.1 (0.62 to 1.58) | 0.88 (0.39 to 1.38) | 0.22 (-0.48 to 0.91) | | |
| IL12 | | | | | |
| Raw score | 9.05 (8.95 to 9.15) | 9.12 (9.02 to 9.22) | -0.07 (-0.22 to 0.07) | 0.291 | 0.884 |
| z-score | -0.05 (-0.24 to 0.14) | 0.1 (-0.1 to 0.29) | -0.14 (-0.41 to 0.13) | | |
| IL5Ralpha | | | | | |
| Raw score | 3.27 (3.18 to 3.36) | 3.35 (3.26 to 3.44) | -0.08 (-0.21 to 0.05) | 0.235 | 0.884 |
| z-score | -0.07 (-0.21 to 0.07) | 0.05 (-0.09 to 0.2) | -0.12 (-0.33 to 0.08) | | |

**Table S3A.** (Continued)

| | | | | | |
|---|---|---|---|---|---|
| JAMB | | | | | |
| Raw score | 8.32 (8.25 to 8.38) | 8.33 (8.26 to 8.4) | -0.01 (-0.11 to 0.09) | 0.810 | 0.966 |
| z-score | -0.06 (-0.3 to 0.18) | -0.02 (-0.27 to 0.23) | -0.04 (-0.39 to 0.31) | | |
| **KYNU** | | | | | |
| Raw score | 8.85 (8.71 to 8.98) | 9.12 (8.98 to 9.26) | -0.28 (-0.47 to -0.08) | **0.005** | 0.166 |
| z-score | -0.41 (-0.61 to -0.21) | 0.01 (-0.2 to 0.22) | -0.42 (-0.71 to -0.13) | | |
| **LAIR2** | | | | | |
| Raw score | 4.72 (4.6 to 4.85) | 4.92 (4.8 to 5.05) | -0.2 (-0.38 to -0.02) | **0.028** | 0.517 |
| z-score | -0.13 (-0.24 to -0.01) | 0.06 (-0.06 to 0.18) | -0.19 (-0.35 to -0.02) | | |
| LAT | | | | | |
| Raw score | 5.73 (5.4 to 6.06) | 5.94 (5.6 to 6.28) | -0.21 (-0.69 to 0.26) | 0.372 | 0.891 |
| z-score | -0.24 (-0.48 to 0) | -0.09 (-0.33 to 0.16) | -0.15 (-0.49 to 0.19) | | |
| LAYN | | | | | |
| Raw score | 5.77 (5.7 to 5.84) | 5.8 (5.72 to 5.87) | -0.03 (-0.13 to 0.07) | 0.607 | 0.891 |
| z-score | 0.04 (-0.19 to 0.28) | 0.13 (-0.11 to 0.38) | -0.09 (-0.43 to 0.25) | | |
| LXN | | | | | |
| Raw score | 2.41 (2.27 to 2.56) | 2.33 (2.18 to 2.47) | 0.09 (-0.12 to 0.29) | 0.394 | 0.891 |
| z-score | 0.65 (0.09 to 1.22) | 0.3 (-0.29 to 0.89) | 0.35 (-0.47 to 1.17) | | |
| MANF | | | | | |
| Raw score | 6.36 (6 to 6.73) | 6.75 (6.37 to 7.13) | -0.39 (-0.92 to 0.14) | 0.152 | 0.884 |
| z-score | -0.26 (-0.58 to 0.06) | 0.08 (-0.26 to 0.41) | -0.34 (-0.8 to 0.13) | | |
| MATN3 | | | | | |
| Raw score | 12.97 (12.92 to 13.03) | 12.96 (12.91 to 13.02) | 0.01 (-0.07 to 0.09) | 0.861 | 0.967 |
| z-score | 0 (-0.29 to 0.3) | -0.03 (-0.34 to 0.28) | 0.04 (-0.39 to 0.47) | | |
| MDGA1 | | | | | |
| Raw score | 5.05 (4.96 to 5.14) | 5.05 (4.95 to 5.14) | 0.01 (-0.12 to 0.13) | 0.932 | 0.995 |
| z-score | -0.04 (-0.18 to 0.09) | -0.05 (-0.19 to 0.08) | 0.01 (-0.18 to 0.2) | | |
| **MSR1** | | | | | |
| Raw score | 6.78 (6.72 to 6.84) | 6.93 (6.87 to 6.99) | -0.15 (-0.23 to -0.06) | **0.001** | 0.077 |
| z-score | -0.28 (-0.4 to -0.15) | 0.04 (-0.09 to 0.17) | -0.32 (-0.5 to -0.14) | | |
| N2DL2 | | | | | |
| Raw score | 3.87 (3.8 to 3.94) | 3.9 (3.83 to 3.97) | -0.03 (-0.13 to 0.07) | 0.521 | 0.891 |
| z-score | 0.03 (-0.2 to 0.26) | 0.14 (-0.1 to 0.37) | -0.11 (-0.43 to 0.22) | | |
| NAAA | | | | | |
| Raw score | 3.74 (3.6 to 3.88) | 3.66 (3.51 to 3.8) | 0.09 (-0.11 to 0.28) | 0.400 | 0.891 |
| z-score | 0.02 (-0.21 to 0.25) | -0.12 (-0.36 to 0.12) | 0.14 (-0.19 to 0.47) | | |
| NBL1 | | | | | |
| Raw score | 5.59 (5.57 to 5.62) | 5.61 (5.58 to 5.64) | -0.01 (-0.05 to 0.03) | 0.459 | 0.891 |
| z-score | -0.18 (-0.44 to 0.08) | -0.04 (-0.3 to 0.23) | -0.14 (-0.52 to 0.24) | | |
| NCAN | | | | | |
| Raw score | 9.8 (9.76 to 9.84) | 9.77 (9.74 to 9.81) | 0.03 (-0.03 to 0.08) | 0.359 | 0.884 |
| z-score | -0.21 (-0.38 to -0.04) | -0.33 (-0.5 to -0.15) | 0.11 (-0.13 to 0.36) | | |
| NCDase | | | | | |
| Raw score | 4.17 (4.1 to 4.24) | 4.14 (4.07 to 4.21) | 0.03 (-0.06 to 0.13) | 0.502 | 0.891 |
| z-score | -0.03 (-0.17 to 0.11) | -0.1 (-0.25 to 0.05) | 0.07 (-0.14 to 0.28) | | |
| NEP | | | | | |
| Raw score | 3.07 (3.01 to 3.13) | 3.05 (2.99 to 3.11) | 0.02 (-0.06 to 0.1) | 0.648 | 0.891 |
| z-score | -0.04 (-0.14 to 0.05) | -0.07 (-0.17 to 0.03) | 0.03 (-0.11 to 0.17) | | |
| NMNAT1 | | | | | |
| Raw score | 4.82 (4.51 to 5.14) | 4.6 (4.27 to 4.93) | 0.22 (-0.23 to 0.68) | 0.334 | 0.884 |
| z-score | 0.47 (0.19 to 0.75) | 0.27 (-0.02 to 0.56) | 0.2 (-0.21 to 0.6) | | |
| NrCAM | | | | | |
| Raw score | 10.12 (10.09 to 10.15) | 10.1 (10.07 to 10.13) | 0.02 (-0.02 to 0.06) | 0.354 | 0.884 |
| z-score | -0.25 (-0.53 to 0.02) | -0.44 (-0.72 to -0.16) | 0.18 (-0.21 to 0.58) | | |
| NRP2 | | | | | |
| Raw score | 8.95 (8.93 to 8.97) | 8.94 (8.92 to 8.97) | 0.01 (-0.03 to 0.04) | 0.663 | 0.891 |
| z-score | -0.25 (-0.49 to -0.01) | -0.33 (-0.58 to -0.08) | 0.08 (-0.27 to 0.42) | | |
| NTRK2 | | | | | |
| Raw score | 7.11 (7.06 to 7.15) | 7.12 (7.07 to 7.16) | -0.01 (-0.07 to 0.05) | 0.759 | 0.962 |
| z-score | -0.37 (-0.66 to -0.09) | -0.31 (-0.6 to -0.01) | -0.06 (-0.48 to 0.35) | | |
| NTRK3 | | | | | |
| Raw score | 7.45 (7.4 to 7.51) | 7.47 (7.42 to 7.53) | -0.02 (-0.09 to 0.06) | 0.666 | 0.891 |
| z-score | -0.3 (-0.54 to -0.05) | -0.22 (-0.48 to 0.04) | -0.08 (-0.44 to 0.28) | | |
| PDGFRalpha | | | | | |
| Raw score | 5.84 (5.78 to 5.9) | 5.85 (5.79 to 5.92) | -0.01 (-0.1 to 0.08) | 0.853 | 0.967 |
| z-score | -0.01 (-0.25 to 0.23) | 0.03 (-0.22 to 0.27) | -0.03 (-0.38 to 0.31) | | |

**Table S3A.** (Continued)

| | | | | | |
|---|---|---|---|---|---|
| PLXNB1 | | | | | |
| Raw score | 1.91 (1.85 to 1.96) | 1.89 (1.83 to 1.94) | 0.02 (-0.05 to 0.1) | 0.577 | 0.891 |
| z-score | -0.19 (-0.4 to 0.02) | -0.28 (-0.49 to -0.06) | 0.09 (-0.22 to 0.38) | | |
| **PLXNB3** | | | | | |
| Raw score | 4.18 (4.09 to 4.27) | 4.33 (4.24 to 4.43) | -0.15 (-0.28 to -0.02) | **0.028** | 0.517 |
| z-score | -0.36 (-0.48 to -0.23) | -0.16 (-0.28 to -0.03) | -0.2 (-0.37 to -0.02) | | |
| PRTG | | | | | |
| Raw score | 7.19 (7.12 to 7.27) | 7.14 (7.06 to 7.22) | 0.05 (-0.06 to 0.16) | 0.349 | 0.884 |
| z-score | -0.03 (-0.26 to 0.2) | -0.19 (-0.43 to 0.05) | 0.16 (-0.18 to 0.49) | | |
| PVR | | | | | |
| Raw score | 8.83 (8.78 to 8.88) | 8.85 (8.8 to 8.89) | -0.02 (-0.09 to 0.05) | 0.599 | 0.891 |
| z-score | -0.15 (-0.32 to 0.02) | -0.08 (-0.26 to 0.1) | -0.06 (-0.31 to 0.18) | | |
| RGMA | | | | | |
| Raw score | 11.04 (10.99 to 11.1) | 11.05 (11 to 11.11) | -0.01 (-0.09 to 0.07) | 0.782 | 0.968 |
| z-score | -0.17 (-0.41 to 0.08) | -0.12 (-0.37 to 0.14) | -0.05 (-0.41 to 0.31) | | |
| RGMB | | | | | |
| Raw score | 6.27 (6.2 to 6.34) | 6.3 (6.23 to 6.38) | -0.03 (-0.13 to 0.07) | 0.525 | 0.891 |
| z-score | -0.07 (-0.35 to 0.2) | 0.05 (-0.23 to 0.34) | -0.13 (-0.53 to 0.27) | | |
| ROBO2 | | | | | |
| Raw score | 6.97 (6.91 to 7.04) | 7.02 (6.96 to 7.09) | -0.05 (-0.14 to 0.04) | 0.290 | 0.884 |
| z-score | -0.3 (-0.53 to -0.07) | -0.12 (-0.36 to 0.11) | -0.18 (-0.5 to 0.15) | | |
| RSPO1 | | | | | |
| Raw score | 1.91 (1.85 to 1.98) | 1.97 (1.9 to 2.04) | -0.05 (-0.15 to 0.04) | 0.277 | 0.884 |
| z-score | -0.24 (-0.46 to -0.02) | -0.06 (-0.3 to 0.17) | -0.18 (-0.5 to 0.14) | | |
| SCARA5 | | | | | |
| Raw score | 9.26 (9.21 to 9.32) | 9.3 (9.24 to 9.35) | -0.03 (-0.11 to 0.05) | 0.441 | 0.891 |
| z-score | -0.09 (-0.36 to 0.19) | 0.07 (-0.22 to 0.36) | -0.16 (-0.56 to 0.24) | | |
| **SCARB2** | | | | | |
| Raw score | 4.12 (4.06 to 4.18) | 4.24 (4.18 to 4.3) | -0.13 (-0.21 to -0.04) | **0.004** | 0.163 |
| z-score | -0.08 (-0.29 to 0.13) | 0.38 (0.16 to 0.6) | -0.46 (-0.77 to -0.16) | | |
| SCARF2 | | | | | |
| Raw score | 7.13 (7.07 to 7.2) | 7.1 (7.04 to 7.17) | 0.03 (-0.07 to 0.12) | 0.545 | 0.891 |
| z-score | -0.1 (-0.36 to 0.16) | -0.22 (-0.49 to 0.06) | 0.12 (-0.26 to 0.5) | | |
| sFRP3 | | | | | |
| Raw score | 3.2 (3.04 to 3.35) | 3.32 (3.15 to 3.48) | -0.12 (-0.34 to 0.1) | 0.295 | 0.884 |
| z-score | -0.28 (-0.47 to -0.09) | -0.13 (-0.33 to 0.06) | -0.14 (-0.42 to 0.13) | | |
| SIGLEC1 | | | | | |
| Raw score | 6.55 (6.46 to 6.64) | 6.64 (6.54 to 6.73) | -0.08 (-0.22 to 0.05) | 0.216 | 0.884 |
| z-score | 0 (-0.19 to 0.19) | 0.17 (-0.03 to 0.37) | -0.17 (-0.45 to 0.1) | | |
| Siglec9 | | | | | |
| Raw score | 5.07 (5.03 to 5.11) | 5.08 (5.03 to 5.12) | -0.01 (-0.07 to 0.05) | 0.768 | 0.962 |
| z-score | -0.12 (-0.26 to 0.03) | -0.09 (-0.23 to 0.06) | -0.03 (-0.23 to 0.17) | | |
| SKR3 | | | | | |
| Raw score | 6.47 (6.42 to 6.52) | 6.5 (6.46 to 6.55) | -0.04 (-0.1 to 0.03) | 0.286 | 0.884 |
| z-score | -0.12 (-0.3 to 0.05) | 0.01 (-0.17 to 0.2) | -0.14 (-0.4 to 0.12) | | |
| SMOC2 | | | | | |
| Raw score | 8.89 (8.82 to 8.96) | 8.92 (8.85 to 9) | -0.03 (-0.14 to 0.07) | 0.544 | 0.891 |
| z-score | -0.01 (-0.23 to 0.21) | 0.09 (-0.14 to 0.32) | -0.1 (-0.42 to 0.22) | | |
| SMPD1 | | | | | |
| Raw score | 4.64 (4.57 to 4.72) | 4.61 (4.53 to 4.68) | 0.04 (-0.07 to 0.15) | 0.481 | 0.891 |
| z-score | 0.15 (-0.08 to 0.37) | 0.03 (-0.2 to 0.27) | 0.12 (-0.21 to 0.44) | | |
| SPOCK1 | | | | | |
| Raw score | 2.82 (2.77 to 2.86) | 2.87 (2.82 to 2.91) | -0.05 (-0.12 to 0.01) | 0.099 | 0.884 |
| z-score | -0.17 (-0.36 to 0.03) | 0.07 (-0.13 to 0.27) | -0.24 (-0.52 to 0.05) | | |
| THY1 | | | | | |
| Raw score | 10.59 (10.55 to 10.64) | 10.58 (10.54 to 10.63) | 0.01 (-0.05 to 0.07) | 0.751 | 0.962 |
| z-score | 0.19 (-0.1 to 0.47) | 0.12 (-0.17 to 0.42) | 0.06 (-0.34 to 0.47) | | |
| TMPRSS5 | | | | | |
| Raw score | 3.3 (3.24 to 3.35) | 3.24 (3.19 to 3.3) | 0.05 (-0.03 to 0.13) | 0.191 | 0.884 |
| z-score | -0.07 (-0.27 to 0.12) | -0.26 (-0.47 to -0.06) | 0.19 (-0.1 to 0.48) | | |
| TNFRSF12A | | | | | |
| Raw score | 5.27 (5.19 to 5.36) | 5.27 (5.18 to 5.36) | 0 (-0.12 to 0.13) | 0.974 | 0.995 |
| z-score | -0.14 (-0.42 to 0.14) | -0.15 (-0.44 to 0.14) | 0.01 (-0.41 to 0.42) | | |
| TNFRSF21 | | | | | |
| Raw score | 8.57 (8.53 to 8.61) | 8.57 (8.54 to 8.61) | 0 (-0.06 to 0.05) | 0.914 | 0.990 |
| z-score | -0.21 (-0.42 to 0) | -0.19 (-0.41 to 0.03) | -0.02 (-0.32 to 0.29) | | |

**Table S3A.** (Continued)

| | | | | | |
|---|---|---|---|---|---|
| TNR | | | | | |
| Raw score | 4.95 (4.87 to 5.02) | 4.99 (4.91 to 5.06) | -0.04 (-0.15 to 0.07) | 0.457 | 0.891 |
| z-score | -0.06 (-0.25 to 0.14) | 0.05 (-0.16 to 0.25) | -0.11 (-0.39 to 0.18) | | |
| UNC5C | | | | | |
| Raw score | 5.04 (4.98 to 5.09) | 5.07 (5.01 to 5.13) | -0.04 (-0.12 to 0.04) | 0.357 | 0.884 |
| z-score | -0.18 (-0.35 to -0.02) | -0.07 (-0.24 to 0.1) | -0.11 (-0.35 to 0.13) | | |
| VWC2 | | | | | |
| Raw score | 6.24 (6.16 to 6.33) | 6.32 (6.23 to 6.4) | -0.07 (-0.19 to 0.05) | 0.229 | 0.884 |
| z-score | -0.09 (-0.31 to 0.13) | 0.1 (-0.13 to 0.33) | -0.19 (-0.51 to 0.12) | | |
| WFIKKN1 | | | | | |
| Raw score | 3.71 (3.65 to 3.76) | 3.78 (3.72 to 3.83) | -0.07 (-0.15 to 0.01) | 0.099 | 0.884 |
| z-score | -0.09 (-0.25 to 0.08) | 0.11 (-0.06 to 0.28) | -0.2 (-0.43 to 0.04) | | |

z Score values indicate how many standard deviations have the post-intervention values changed with respect to the baseline mean and standard deviation. E.g., a 0.50 z Score means that the mean value at post-intervention is 0.50 standard deviations higher than the mean value at baseline, indicating a positive change, with negative values indicating the opposite. Values are expressed as mean (95% CI). Analyses were adjusted for baseline values. N=81 participants. Raw scores values are expressed in Normalized Protein eXpression (NPX) units. This means that a high NPX value equals a high protein concentration. Because NPX is in a log2 scale, a 1 NPX difference means a doubling of protein concentration. If needed NPX values can be converted into linear scale: 2NPX= linear NPX. Abbreviations: $p_{FDR}$= adjusted for multiple comparisons using false discovery rate.

**Table S4A.** ELISA assay technique validation

| | $N_{all}$ | $N_I$ | Intervention group | $N_c$ | Control group | Differences between groups | *p* |
|---|---|---|---|---|---|---|---|
| **Intention-to-treat** | | | | | | | |
| MSR1 (pg/ml) | | | | | | | |
| Raw score | 83 | 45 | 249.36 (223.72, 275.00) | 38 | 293.88 (265.98, 321.78) | -44.52 (-82.42, -6.63) | **0.022** |
| z-score | | | -0.14 (-0.30, 0.03) | | 0.15 (-0.03, 0.33) | -0.29 (-0.53, -0.04) | |
| PLXNB3 (pg/ml) | | | | | | | |
| Raw score | 81 | 45 | 2073.95 (1715.68, 2432.22) | 36 | 2152.41 (1751.42, 2553.39) | -78.46 (-618.71, 461.79) | 0.773 |
| z-score | | | -0.01 (-0.07, 0.06) | | 0.01 (-0.06, 0.08) | -0.01 (-0.11, 0.08) | |
| KYNU (ng/ml) | | | | | | | |
| Raw score | 87 | 48 | 74.98 (32.52, 117.45) | 39 | 94.27 (47.13, 141.41) | -19.29 (-82.90, 44.33) | 0.548 |
| z-score | | | 0.06 (-0.22, 0.34) | | 0.18 (-0.13, 0.50) | -0.13 (-0.55, 0.29) | |

z Score values indicate how many standard deviations have the post-intervention values changed with respect to the baseline mean and standard deviation. E.g., a 0.50 z Score means that the mean value at post-intervention is 0.50 standard deviations higher than the mean value at baseline, indicating a positive change, with negative values indicating the opposite. Values are expressed as mean (95% CI). Analyses were adjusted for baseline values.

**Table S5A.** Intention-to-treat effects of the ActiveBrains intervention on raw and z Score post-intervention novel blood-circulating biomarkers identified based on previous literature.

| | | | Mean (95% CI) | | | | |
|---|---|---|---|---|---|---|---|
| | $N_{all}$ | $N_I$ | Intervention group | $N_c$ | Control group | Differences between groups | *p* |
| BDNF (ng/ml) | | | | | | | |
| Raw score | 85 | 47 | 3.84 (2.97, 4.71) | 38 | 2.91 (1.95, 3.88) | 0.92 (-0.38, 2.23) | 0.162 |
| z-score | | | -0.16 (-0.36, 0.03) | | -0.37 (-0.59, -0.15) | 0.21 (-0.09, 0.50) | |
| BHB (ng/µl) | | | | | | | |
| Raw score | 86 | 47 | 31.61 (28.23, 34.99) | 39 | 32.33 (28.62, 36.05) | -0.72 (-5.76, 4.32) | 0.777 |
| z-score | | | -0.30 (-0.68, 0.08) | | -0.22 (-0.64, 0.20) | -0.08 (-0.65, 0.48) | |
| CTSB (µg/l) | | | | | | | |
| Raw score | 86 | 47 | 56.01 (51.16, 60.86) | 39 | 59.15 (53.82, 64.48) | -3.14 (-10.41, 4.14) | 0.393 |
| z-score | | | -0.08 (-0.31, 0.15) | | 0.07 (-0.18, 0.33) | -0.15 (-0.50, 0.20) | |
| Kynurenine (ng/ml) | | | | | | | |
| Raw score | 87 | 48 | 1134.93 (1055.48, 1214.37) | 39 | 1079.54 (991.40, 1167.67) | -55.39 ( -174.05, 63.28) | 0.356 |
| z-score | | | 0.32 (0.13, 0.51) | | 0.19 (-0.02, 0.40) | 0.13 (-0.15, 0.41) | |
| FGF21 (pg/ml) | | | | | | | |
| Raw score | 84 | 45 | 79.62 (60.37, 98.87) | 39 | 103.92 (83.24, 124.61) | -24.30 (-52.59, 3.98) | 0.091 |
| z-score | | | -0.10 (-0.27, 0.08) | | 0.12 (-0.06, 0.31) | -0.22 (-0.47, 0.03) | |
| sVCAM-1 (µg/l) | | | | | | | |
| Raw score | 86 | 47 | 1268.18 (1200.73, 1335.63) | 39 | 1307.57 (1233.52, 1381.62) | -39.39 (-139.58, 60.80) | 0.436 |
| z-score | | | 0.06 (-0.18, 0.31) | | 0.21 (-0.06, 0.48) | -0.14 (-0.51, 0.22) | |

z Score values indicate how many standard deviations have the post-intervention values changed with respect to the baseline mean and standard deviation. E.g., a 0.50 z Score means that the mean value at post-intervention is 0.50 standard deviations higher than the mean value at baseline, indicating a positive change, with negative values indicating the opposite. Values are expressed as mean (95% CI). Analyses were adjusted for baseline values. Abbreviations: BHB= B-hydroxybutyrate, CTSB= Cathepsin B, FGF21= Fibroblast growth factor 21, sVCAM-1= vascular cell adhesion molecule-1.

**Table S6A.** Intention-to-treat effects of the ActiveBrains intervention on raw and z Score post-intervention neurology-related proteins (n=87).

| | Intervention group | Control group | Differences between groups | *p* | $p_{FDR}$ |
|---|---|---|---|---|---|
| ADAM22 | | | | | |
| Raw score | 5.00 (4.92 to 5.07) | 4.97 (4.89 to 5.06) | 0.02 (-0.09 to 0.13) | 0.685 | 0.998 |
| z-score | 0.14 (-0.10 to 0.38) | 0.06 (-0.20 to 0.33) | 0.07 (-0.28 to 0.43) | | |
| ADAM23 | | | | | |
| Raw score | 4.97 (4.90 to 5.04) | 4.91 (4.83 to 4.99) | 0.06 (-0.05 to 0.16) | 0.276 | 0.935 |
| z-score | -0.01 (-0.22 to 0.20) | -0.19 (-0.42 to 0.05) | 0.17 (-0.14 to 0.49) | | |
| Alpha2MRAP | | | | | |
| Raw score | 8.02 (7.86 to 8.18) | 8.03 (7.85 to 8.21) | 0.00 (-0.24 to 0.23) | 0.970 | 0.998 |
| z-score | -0.17 (-0.34 to 0.00) | -0.16 (-0.35 to 0.03) | 0.00 (-0.26 to 0.25) | | |
| BCAN | | | | | |
| Raw score | 5.05 (4.99 to 5.11) | 5.09 (5.02 to 5.15) | -0.04 (-0.13 to 0.05) | 0.396 | 0.935 |
| z-score | -0.3 (-0.5 to -0.11) | -0.18 (-0.39 to 0.03) | -0.12 (-0.41 to 0.16) | | |
| BetaNGF | | | | | |
| Raw score | 1.28 (1.24 to 1.32) | 1.29 (1.25 to 1.33) | -0.01 (-0.06 to 0.05) | 0.799 | 0.998 |
| z-score | -0.19 (-0.37 to -0.02) | -0.16 (-0.35 to 0.03) | -0.03 (-0.29 to 0.22) | | |
| BMP4 | | | | | |
| Raw score | 4.46 (4.39 to 4.53) | 4.45 (4.37 to 4.53) | 0.01 (-0.10 to 0.12) | 0.880 | 0.998 |
| z-score | 0.05 (-0.20 to 0.30) | 0.02 (-0.25 to 0.29) | 0.03 (-0.34 to 0.40) | | |
| CADM3 | | | | | |
| Raw score | 4.83 (4.75 to 4.91) | 4.82 (4.74 to 4.91) | 0.01 (-0.11 to 0.12) | 0.920 | 0.998 |
| z-score | -0.12 (-0.29 to 0.06) | -0.13 (-0.32 to 0.06) | 0.01 (-0.25 to 0.27) | | |
| CD200 | | | | | |
| Raw score | 7.05 (6.99 to 7.11) | 7.05 (6.98 to 7.11) | 0.00 (-0.08 to 0.09) | 0.903 | 0.998 |
| z-score | -0.07 (-0.29 to 0.16) | -0.09 (-0.34 to 0.16) | 0.02 (-0.31 to 0.36) | | |
| CD200R1 | | | | | |
| Raw score | 5.15 (5.10 to 5.20) | 5.16 (5.11 to 5.22) | -0.01 (-0.09 to 0.06) | 0.674 | 0.998 |
| z-score | -0.11 (-0.27 to 0.05) | -0.06 (-0.23 to 0.12) | -0.05 (-0.29 to 0.19) | | |
| CD38 | | | | | |
| Raw score | 5.96 (5.89 to 6.02) | 5.99 (5.92 to 6.06) | -0.04 (-0.13 to 0.06) | 0.425 | 0.935 |
| z-score | -0.24 (-0.52 to 0.03) | -0.08 (-0.38 to 0.23) | -0.17 (-0.58 to 0.24) | | |
| CDH3 | | | | | |
| Raw score | 8.07 (8.00 to 8.14) | 8.12 (8.04 to 8.19) | -0.05 (-0.15 to 0.05) | 0.342 | 0.935 |
| z-score | -0.06 (-0.28 to 0.15) | 0.09 (-0.15 to 0.33) | -0.16 (-0.48 to 0.17) | | |
| CDH6 | | | | | |
| Raw score | 5.39 (5.35 to 5.44) | 5.38 (5.33 to 5.43) | 0.01 (-0.06 to 0.08) | 0.781 | 0.998 |
| z-score | -0.1 (-0.35 to 0.15) | -0.15 (-0.43 to 0.12) | 0.05 (-0.32 to 0.43) | | |
| CLEC10A | | | | | |
| Raw score | 5.62 (5.54 to 5.7) | 5.69 (5.6 to 5.78) | -0.07 (-0.2 to 0.05) | 0.231 | 0.935 |
| z-score | -0.12 (-0.33 to 0.09) | 0.07 (-0.16 to 0.30) | -0.19 (-0.5 to 0.12) | | |
| CLEC1B | | | | | |
| Raw score | 9.93 (9.67 to 10.19) | 9.9 (9.62 to 10.19) | 0.03 (-0.36 to 0.42) | 0.892 | 0.998 |
| z-score | -0.2 (-0.43 to 0.02) | -0.23 (-0.48 to 0.02) | 0.02 (-0.32 to 0.36) | | |
| CLM1 | | | | | |
| Raw score | 6.31 (6.23 to 6.39) | 6.31 (6.22 to 6.4) | 0.00 (-0.12 to 0.12) | 0.998 | 0.998 |
| z-score | -0.03 (-0.16 to 0.11) | -0.03 (-0.18 to 0.13) | 0.00 (-0.21 to 0.21) | | |
| CLM6 | | | | | |
| Raw score | 5.8 (5.76 to 5.84) | 5.83 (5.78 to 5.87) | -0.03 (-0.08 to 0.03) | 0.380 | 0.935 |
| z-score | -0.13 (-0.28 to 0.03) | -0.03 (-0.20 to 0.14) | -0.10 (-0.33 to 0.13) | | |
| CNTN5 | | | | | |
| Raw score | 6.19 (6.12 to 6.27) | 6.16 (6.07 to 6.24) | 0.04 (-0.08 to 0.15) | 0.552 | 0.992 |
| z-score | -0.23 (-0.44 to -0.03) | -0.32 (-0.55 to -0.10) | 0.09 (-0.21 to 0.4) | | |
| CPA2 | | | | | |
| Raw score | 9.93 (9.81 to 10.05) | 10.10 (9.97 to 10.23) | -0.17 (-0.35 to 0.01) | 0.067 | 0.914 |
| z-score | -0.06 (-0.25 to 0.12) | 0.20 (0.00 to 0.41) | -0.26 (-0.55 to 0.02) | | |
| CPM | | | | | |
| Raw score | 7.08 (7.04 to 7.12) | 7.05 (7.00 to 7.10) | 0.03 (-0.04 to 0.09) | 0.379 | 0.935 |
| z-score | 0.07 (-0.11 to 0.25) | -0.05 (-0.25 to 0.15) | 0.12 (-0.15 to 0.39) | | |
| CRTAM | | | | | |
| Raw score | 5.95 (5.88 to 6.02) | 5.95 (5.88 to 6.03) | 0.00 (-0.11 to 0.10) | 0.928 | 0.998 |
| z-score | -0.16 (-0.29 to -0.03) | -0.15 (-0.29 to -0.01) | -0.01 (-0.20 to 0.18) | | |
| CTSC | | | | | |
| Raw score | 2.98 (2.89 to 3.07) | 2.86 (2.76 to 2.96) | 0.12 (-0.02 to 0.25) | 0.098 | 0.935 |
| z-score | 0.04 (-0.15 to 0.22) | -0.20 (-0.40 to 0.01) | 0.23 (-0.04 to 0.51) | | |

**Table S6A.** (Continued)

| | | | | | |
|---|---|---|---|---|---|
| CTSS | | | | | |
| Raw score | 5.67 (5.63 to 5.72) | 5.61 (5.56 to 5.66) | 0.06 (0.00 to 0.13) | 0.070 | 0.914 |
| z-score | 0.23 (0.00 to 0.46) | -0.09 (-0.35 to 0.16) | 0.32 (-0.03 to 0.66) | | |
| DDR1 | | | | | |
| Raw score | 7.74 (7.7 to 7.78) | 7.75 (7.70 to 7.79) | 0.00 (-0.07 to 0.06) | 0.876 | 0.998 |
| z-score | -0.16 (-0.39 to 0.07) | -0.13 (-0.38 to 0.12) | -0.03 (-0.37 to 0.31) | | |
| Dkk4 | | | | | |
| Raw score | 2.38 (2.27 to 2.49) | 2.50 (2.38 to 2.63) | -0.13 (-0.3 to 0.04) | 0.147 | 0.935 |
| z-score | -0.08 (-0.4 to 0.24) | 0.28 (-0.08 to 0.64) | -0.36 (-0.84 to 0.13) | | |
| DRAXIN | | | | | |
| Raw score | 4.24 (4.15 to 4.33) | 4.24 (4.14 to 4.34) | 0 (-0.13 to 0.13) | 0.993 | 0.998 |
| z-score | -0.08 (-0.26 to 0.10) | -0.08 (-0.28 to 0.12) | 0 (-0.28 to 0.27) | | |
| EDA2R | | | | | |
| Raw score | 3.75 (3.68 to 3.82) | 3.77 (3.69 to 3.85) | -0.01 (-0.12 to 0.09) | 0.774 | 0.998 |
| z-score | 0.22 (0.01 to 0.43) | 0.26 (0.03 to 0.50) | -0.04 (-0.36 to 0.27) | | |
| EFNA4 | | | | | |
| Raw score | 3.53 (3.48 to 3.58) | 3.57 (3.51 to 3.62) | -0.03 (-0.11 to 0.04) | 0.368 | 0.935 |
| z-score | -0.06 (-0.31 to 0.19) | 0.11 (-0.16 to 0.38) | -0.17 (-0.54 to 0.20) | | |
| EPHB6 | | | | | |
| Raw score | 4.93 (4.88 to 4.98) | 4.92 (4.87 to 4.98) | 0.00 (-0.08 to 0.08) | 0.937 | 0.998 |
| z-score | -0.21 (-0.40 to -0.01) | -0.22 (-0.43 to 0.00) | 0.01 (-0.28 to 0.30) | | |
| EZR | | | | | |
| Raw score | 5.14 (5.08 to 5.21) | 5.16 (5.09 to 5.23) | -0.01 (-0.11 to 0.09) | 0.773 | 0.998 |
| z-score | 0.14 (-0.10 to 0.38) | 0.2 (-0.07 to 0.46) | -0.05 (-0.42 to 0.31) | | |
| FcRL2 | | | | | |
| Raw score | 5.37 (5.31 to 5.43) | 5.43 (5.37 to 5.5) | -0.07 (-0.16 to 0.02) | 0.132 | 0.935 |
| z-score | -0.19 (-0.33 to -0.04) | -0.02 (-0.18 to 0.14) | -0.17 (-0.39 to 0.05) | | |
| FLRT2 | | | | | |
| Raw score | 3.24 (3.17 to 3.3) | 3.25 (3.18 to 3.32) | -0.01 (-0.11 to 0.08) | 0.763 | 0.998 |
| z-score | -0.05 (-0.39 to 0.3) | 0.03 (-0.35 to 0.41) | -0.08 (-0.59 to 0.44) | | |
| gal8 | | | | | |
| Raw score | 5.75 (5.64 to 5.87) | 5.74 (5.61 to 5.86) | 0.01 (-0.16 to 0.19) | 0.859 | 0.998 |
| z-score | -0.22 (-0.35 to -0.10) | -0.24 (-0.38 to -0.10) | 0.02 (-0.17 to 0.20) | | |
| GCP5 | | | | | |
| Raw score | 5.35 (5.22 to 5.49) | 5.25 (5.10 to 5.40) | 0.11 (-0.10 to 0.31) | 0.302 | 0.935 |
| z-score | -0.15 (-0.36 to 0.06) | -0.31 (-0.55 to -0.08) | 0.16 (-0.15 to 0.48) | | |
| GCSF | | | | | |
| Raw score | 3.62 (3.48 to 3.76) | 3.72 (3.56 to 3.88) | -0.10 (-0.31 to 0.11) | 0.359 | 0.935 |
| z-score | -0.23 (-0.47 to 0.01) | -0.06 (-0.33 to 0.20) | -0.16 (-0.52 to 0.19) | | |
| GDF8 | | | | | |
| Raw score | 4.27 (4.18 to 4.37) | 4.22 (4.11 to 4.32) | 0.05 (-0.09 to 0.20) | 0.455 | 0.935 |
| z-score | 0.14 (-0.11 to 0.40) | 0 (-0.28 to 0.28) | 0.14 (-0.23 to 0.52) | | |
| GDNF | | | | | |
| Raw score | 1.38 (1.30 to 1.46) | 1.36 (1.27 to 1.45) | 0.02 (-0.11 to 0.14) | 0.776 | 0.998 |
| z-score | 0.10 (-0.24 to 0.45) | 0.03 (-0.35 to 0.41) | 0.07 (-0.44 to 0.58) | | |
| GDNFRalpha 3 | | | | | |
| Raw score | 5.55 (5.49 to 5.60) | 5.54 (5.47 to 5.60) | 0.01 (-0.07 to 0.10) | 0.757 | 0.998 |
| z-score | -0.03 (-0.24 to 0.18) | -0.08 (-0.32 to 0.15) | 0.05 (-0.27 to 0.37) | | |
| GFRalpha1 | | | | | |
| Raw score | 7.56 (7.51 to 7.62) | 7.56 (7.49 to 7.62) | 0.01 (-0.07 to 0.09) | 0.843 | 0.998 |
| z-score | -0.06 (-0.29 to 0.16) | -0.10 (-0.35 to 0.15) | 0.03 (-0.30 to 0.37) | | |
| GMCSFRalpha | | | | | |
| Raw score | 6.10 (6.04 to 6.16) | 6.14 (6.07 to 6.20) | -0.03 (-0.12 to 0.06) | 0.462 | 0.935 |
| z-score | -0.10 (-0.18 to -0.01) | -0.05 (-0.14 to 0.04) | -0.05 (-0.17 to 0.08) | | |
| GZMA | | | | | |
| Raw score | 6.38 (6.27 to 6.49) | 6.45 (6.33 to 6.58) | -0.07 (-0.24 to 0.09) | 0.384 | 0.935 |
| z-score | -0.07 (-0.28 to 0.13) | 0.06 (-0.17 to 0.28) | -0.13 (-0.44 to 0.17) | | |
| HAGH | | | | | |
| Raw score | 6.28 (5.98 to 6.58) | 6.14 (5.81 to 6.48) | 0.14 (-0.32 to 0.59) | 0.556 | 0.992 |
| z-score | 1.1 (0.64 to 1.55) | 0.89 (0.38 to 1.40) | 0.2 (-0.48 to 0.89) | | |
| IL12 | | | | | |
| Raw score | 9.08 (8.99 to 9.17) | 9.14 (9.03 to 9.24) | -0.06 (-0.19 to 0.08) | 0.422 | 0.935 |
| z-score | -0.02 (-0.20 to 0.16) | 0.09 (-0.11 to 0.28) | -0.11 (-0.37 to 0.16) | | |

**Table S6A.** (Continued)

| | | | | | |
|---|---|---|---|---|---|
| IL5Ralpha | | | | | |
| Raw score | 3.31 (3.22 to 3.39) | 3.35 (3.26 to 3.44) | -0.05 (-0.17 to 0.08) | 0.452 | 0.935 |
| z-score | -0.02 (-0.15 to 0.11) | 0.05 (-0.09 to 0.2) | -0.07 (-0.27 to 0.12) | | |
| JAMB | | | | | |
| Raw score | 8.33 (8.26 to 8.39) | 8.32 (8.25 to 8.4) | 0.00 (-0.10 to 0.10) | 0.966 | 0.998 |
| z-score | 0.01 (-0.23 to 0.24) | 0 (-0.26 to 0.26) | 0.01 (-0.34 to 0.36) | | |
| **KYNU** | | | | | |
| Raw score | 8.89 (8.77 to 9.01) | 9.12 (8.98 to 9.26) | -0.23 (-0.42 to -0.05) | **0.015** | 0.532 |
| z-score | -0.36 (-0.55 to -0.16) | 0.01 (-0.21 to 0.22) | -0.37 (-0.66 to -0.07) | | |
| **LAIR2** | | | | | |
| Raw score | 4.81 (4.70 to 4.93) | 4.97 (4.84 to 5.10) | -0.16 (-0.33 to 0.01) | ***0.070*** | 0.914 |
| z-score | -0.09 (-0.20 to 0.01) | 0.05 (-0.06 to 0.17) | -0.14 (-0.30 to 0.01) | | |
| LAT | | | | | |
| Raw score | 5.83 (5.52 to 6.14) | 5.94 (5.60 to 6.29) | -0.12 (-0.58 to 0.35) | 0.627 | 0.998 |
| z-score | -0.17 (-0.40 to 0.06) | -0.08 (-0.34 to 0.17) | -0.08 (-0.42 to 0.26) | | |
| LAYN | | | | | |
| Raw score | 5.77 (5.70 to 5.83) | 5.8 (5.72 to 5.87) | -0.03 (-0.13 to 0.07) | 0.569 | 0.995 |
| z-score | 0.05 (-0.17 to 0.27) | 0.14 (-0.10 to 0.39) | -0.10 (-0.44 to 0.24) | | |
| LXN | | | | | |
| Raw score | 2.41 (2.28 to 2.54) | 2.33 (2.18 to 2.47) | 0.08 (-0.11 to 0.27) | 0.405 | 0.935 |
| z-score | 0.64 (0.11 to 1.18) | 0.31 (-0.28 to 0.9) | 0.33 (-0.46 to 1.13) | | |
| MANF | | | | | |
| Raw score | 6.47 (6.13 to 6.82) | 6.75 (6.37 to 7.14) | -0.28 (-0.8 to 0.24) | 0.289 | 0.935 |
| z-score | -0.16 (-0.46 to 0.14) | 0.08 (-0.25 to 0.42) | -0.24 (-0.69 to 0.21) | | |
| MATN3 | | | | | |
| Raw score | 12.98 (12.93 to 13.03) | 12.96 (12.9 to 13.02) | 0.02 (-0.06 to 0.1) | 0.636 | 0.998 |
| z-score | 0.09 (-0.2 to 0.38) | -0.02 (-0.34 to 0.3) | 0.10 (-0.34 to 0.55) | | |
| MDGA1 | | | | | |
| Raw score | 5.04 (4.95 to 5.12) | 5.03 (4.93 to 5.12) | 0.01 (-0.12 to 0.13) | 0.895 | 0.998 |
| z-score | -0.04 (-0.16 to 0.09) | -0.05 (-0.19 to 0.09) | 0.01 (-0.17 to 0.20) | | |
| **MSR1** | | | | | |
| Raw score | 6.83 (6.77 to 6.89) | 6.95 (6.88 to 7.01) | -0.11 (-0.2 to -0.03) | **0.012** | 0.532 |
| z-score | -0.21 (-0.34 to -0.08) | 0.04 (-0.10 to 0.18) | -0.25 (-0.44 to -0.06) | | |
| N2DL2 | | | | | |
| Raw score | 3.88 (3.81 to 3.94) | 3.9 (3.82 to 3.97) | -0.02 (-0.12 to 0.08) | 0.714 | 0.998 |
| z-score | 0.08 (-0.14 to 0.30) | 0.14 (-0.10 to 0.38) | -0.06 (-0.39 to 0.27) | | |
| NAAA | | | | | |
| Raw score | 3.77 (3.63 to 3.9) | 3.66 (3.51 to 3.8) | 0.11 (-0.09 to 0.3) | 0.274 | 0.935 |
| z-score | 0.06 (-0.16 to 0.29) | -0.12 (-0.37 to 0.13) | 0.19 (-0.15 to 0.52) | | |
| NBL1 | | | | | |
| Raw score | 5.60 (5.57 to 5.62) | 5.61 (5.58 to 5.64) | -0.01 (-0.05 to 0.03) | 0.478 | 0.942 |
| z-score | -0.16 (-0.41 to 0.09) | -0.03 (-0.30 to 0.25) | -0.14 (-0.52 to 0.24) | | |
| NCAN | | | | | |
| Raw score | 9.81 (9.78 to 9.84) | 9.78 (9.74 to 9.82) | 0.03 (-0.02 to 0.08) | 0.219 | 0.935 |
| z-score | -0.18 (-0.34 to -0.02) | -0.33 (-0.51 to -0.15) | 0.15 (-0.09 to 0.39) | | |
| NCDase | | | | | |
| Raw score | 4.18 (4.12 to 4.25) | 4.12 (4.05 to 4.19) | 0.06 (-0.03 to 0.16) | 0.184 | 0.935 |
| z-score | 0.04 (-0.10 to 0.19) | -0.1 (-0.25 to 0.06) | 0.14 (-0.07 to 0.35) | | |
| NEP | | | | | |
| Raw score | 3.08 (3.02 to 3.14) | 3.05 (2.99 to 3.11) | 0.03 (-0.05 to 0.12) | 0.435 | 0.935 |
| z-score | -0.02 (-0.11 to 0.07) | -0.07 (-0.18 to 0.03) | 0.06 (-0.09 to 0.20) | | |
| NMNAT1 | | | | | |
| Raw score | 4.80 (4.51 to 5.10) | 4.6 (4.27 to 4.92) | 0.21 (-0.23 to 0.64) | 0.344 | 0.935 |
| z-score | 0.48 (0.22 to 0.75) | 0.29 (0.00 to 0.59) | 0.19 (-0.21 to 0.59) | | |
| NrCAM | | | | | |
| Raw score | 10.13 (10.11 to 10.16) | 10.1 (10.07 to 10.13) | 0.03 (-0.01 to 0.08) | 0.149 | 0.935 |
| z-score | -0.14 (-0.41 to 0.13) | -0.44 (-0.74 to -0.14) | 0.3 (-0.11 to 0.70) | | |
| NRP2 | | | | | |
| Raw score | 8.96 (8.93 to 8.98) | 8.94 (8.92 to 8.97) | 0.01 (-0.02 to 0.05) | 0.448 | 0.935 |
| z-score | -0.18 (-0.42 to 0.06) | -0.32 (-0.59 to -0.05) | 0.14 (-0.22 to 0.50) | | |
| NTRK2 | | | | | |
| Raw score | 7.12 (7.08 to 7.16) | 7.12 (7.07 to 7.16) | 0.00 (-0.06 to 0.07) | 0.906 | 0.998 |
| z-score | -0.27 (-0.54 to 0.01) | -0.29 (-0.6 to 0.01) | 0.02 (-0.39 to 0.44) | | |

**Table S6A.** (Continued)

| | | | | | |
|---|---|---|---|---|---|
| NTRK3 | | | | | |
| Raw score | 7.47 (7.42 to 7.52) | 7.47 (7.41 to 7.52) | 0.00 (-0.07 to 0.08) | 0.908 | 0.998 |
| z-score | -0.18 (-0.43 to 0.06) | -0.21 (-0.47 to 0.06) | 0.02 (-0.34 to 0.38) | | |
| PDGFRalpha | | | | | |
| Raw score | 5.86 (5.8 to 5.91) | 5.85 (5.79 to 5.92) | 0.00 (-0.08 to 0.09) | 0.945 | 0.998 |
| z-score | 0.04 (-0.19 to 0.28) | 0.03 (-0.23 to 0.29) | 0.01 (-0.34 to 0.37) | | |
| PLXNB1 | | | | | |
| Raw score | 1.93 (1.88 to 1.99) | 1.89 (1.83 to 1.95) | 0.05 (-0.03 to 0.13) | 0.251 | 0.935 |
| z-score | -0.09 (-0.31 to 0.13) | -0.28 (-0.53 to -0.04) | 0.19 (-0.14 to 0.52) | | |
| **PLXNB3** | | | | | |
| Raw score | 4.21 (4.12 to 4.3) | 4.33 (4.23 to 4.43) | -0.12 (-0.25 to 0.01) | ***0.068*** | 0.914 |
| z-score | -0.31 (-0.43 to -0.19) | -0.14 (-0.28 to -0.01) | -0.17 (-0.35 to 0.01) | | |
| PRTG | | | | | |
| Raw score | 7.19 (7.12 to 7.26) | 7.13 (7.06 to 7.21) | 0.06 (-0.05 to 0.16) | 0.288 | 0.935 |
| z-score | -0.01 (-0.23 to 0.21) | -0.19 (-0.43 to 0.06) | 0.18 (-0.15 to 0.50) | | |
| PVR | | | | | |
| Raw score | 8.84 (8.80 to 8.89) | 8.84 (8.79 to 8.90) | 0 (-0.07 to 0.07) | 0.997 | 0.998 |
| z-score | -0.08 (-0.25 to 0.09) | -0.08 (-0.27 to 0.11) | 0 (-0.26 to 0.26) | | |
| RGMA | | | | | |
| Raw score | 11.05 (11.00 to 11.11) | 11.05 (10.99 to 11.11) | 0.00 (-0.07 to 0.08) | 0.934 | 0.998 |
| z-score | -0.07 (-0.29 to 0.15) | -0.09 (-0.33 to 0.16) | 0.01 (-0.32 to 0.35) | | |
| RGMB | | | | | |
| Raw score | 6.27 (6.21 to 6.34) | 6.3 (6.23 to 6.38) | -0.03 (-0.13 to 0.07) | 0.536 | 0.992 |
| z-score | -0.06 (-0.32 to 0.20) | 0.06 (-0.22 to 0.35) | -0.12 (-0.51 to 0.26) | | |
| ROBO2 | | | | | |
| Raw score | 6.98 (6.92 to 7.04) | 7.02 (6.95 to 7.09) | -0.04 (-0.13 to 0.06) | 0.425 | 0.935 |
| z-score | -0.23 (-0.45 to 0.00) | -0.09 (-0.34 to 0.16) | -0.14 (-0.47 to 0.20) | | |
| RSPO1 | | | | | |
| Raw score | 1.93 (1.86 to 2.00) | 1.97 (1.89 to 2.04) | -0.04 (-0.14 to 0.06) | 0.445 | 0.935 |
| z-score | -0.18 (-0.40 to 0.04) | -0.06 (-0.30 to 0.18) | -0.13 (-0.45 to 0.20) | | |
| SCARA5 | | | | | |
| Raw score | 9.27 (9.21 to 9.32) | 9.29 (9.23 to 9.35) | -0.03 (-0.11 to 0.05) | 0.499 | 0.945 |
| z-score | -0.04 (-0.3 to 0.23) | 0.1 (-0.20 to 0.40) | -0.14 (-0.54 to 0.27) | | |
| **SCARB2** | | | | | |
| Raw score | 4.14 (4.09 to 4.2) | 4.24 (4.18 to 4.31) | -0.1 (-0.19 to -0.02) | **0.018** | 0.532 |
| z-score | 0 (-0.21 to 0.21) | 0.38 (0.15 to 0.61) | -0.38 (-0.69 to -0.07) | | |
| SCARF2 | | | | | |
| Raw score | 7.13 (7.07 to 7.20) | 7.1 (7.03 to 7.17) | 0.03 (-0.06 to 0.13) | 0.486 | 0.942 |
| z-score | -0.07 (-0.32 to 0.18) | -0.2 (-0.48 to 0.08) | 0.13 (-0.24 to 0.51) | | |
| sFRP3 | | | | | |
| Raw score | 3.22 (3.08 to 3.37) | 3.31 (3.14 to 3.47) | -0.08 (-0.31 to 0.14) | 0.455 | 0.935 |
| z-score | -0.23 (-0.42 to -0.04) | -0.12 (-0.33 to 0.09) | -0.11 (-0.39 to 0.17) | | |
| SIGLEC1 | | | | | |
| Raw score | 6.57 (6.49 to 6.66) | 6.65 (6.56 to 6.74) | -0.07 (-0.20 to 0.05) | 0.238 | 0.935 |
| z-score | 0.01 (-0.17 to 0.19) | 0.17 (-0.03 to 0.37) | -0.16 (-0.43 to 0.11) | | |
| Siglec9 | | | | | |
| Raw score | 5.09 (5.05 to 5.13) | 5.09 (5.04 to 5.13) | 0.01 (-0.06 to 0.07) | 0.856 | 0.998 |
| z-score | -0.07 (-0.2 to 0.07) | -0.09 (-0.24 to 0.06) | 0.02 (-0.19 to 0.22) | | |
| SKR3 | | | | | |
| Raw score | 6.50 (6.45 to 6.55) | 6.50 (6.45 to 6.56) | -0.01 (-0.08 to 0.07) | 0.865 | 0.998 |
| z-score | 0.00 (-0.20 to 0.19) | 0.02 (-0.19 to 0.24) | -0.03 (-0.31 to 0.26) | | |
| SMOC2 | | | | | |
| Raw score | 8.92 (8.85 to 8.98) | 8.93 (8.85 to 9.00) | -0.01 (-0.12 to 0.10) | 0.847 | 0.998 |
| z-score | 0.06 (-0.16 to 0.28) | 0.09 (-0.15 to 0.33) | -0.03 (-0.36 to 0.30) | | |
| SMPD1 | | | | | |
| Raw score | 4.68 (4.61 to 4.74) | 4.61 (4.54 to 4.69) | 0.06 (-0.04 to 0.17) | 0.231 | 0.935 |
| z-score | 0.22 (0.00 to 0.43) | 0.02 (-0.22 to 0.26) | 0.19 (-0.13 to 0.52) | | |
| SPOCK1 | | | | | |
| Raw score | 2.82 (2.78 to 2.86) | 2.87 (2.82 to 2.92) | -0.05 (-0.11 to 0.02) | 0.139 | 0.935 |
| z-score | -0.14 (-0.33 to 0.06) | 0.09 (-0.13 to 0.30) | -0.22 (-0.52 to 0.07) | | |
| THY1 | | | | | |
| Raw score | 10.60 (10.56 to 10.64) | 10.58 (10.54 to 10.63) | 0.02 (-0.04 to 0.08) | 0.580 | 0.995 |
| z-score | 0.25 (-0.02 to 0.52) | 0.14 (-0.16 to 0.44) | 0.11 (-0.29 to 0.52) | | |

**Table S6A.** (Continued)

| | | | | | |
|---|---|---|---|---|---|
| TMPRSS5 | | | | | |
| Raw score | 3.31 (3.26 to 3.36) | 3.25 (3.19 to 3.3) | 0.06 (-0.01 to 0.14) | 0.115 | 0.935 |
| z-score | -0.04 (-0.22 to 0.14) | -0.26 (-0.47 to -0.06) | 0.22 (-0.06 to 0.5) | | |
| TNFRSF12A | | | | | |
| Raw score | 5.27 (5.2 to 5.35) | 5.27 (5.18 to 5.36) | 0 (-0.12 to 0.12) | 0.976 | 0.998 |
| z-score | -0.15 (-0.42 to 0.12) | -0.16 (-0.45 to 0.14) | 0.01 (-0.4 to 0.41) | | |
| TNFRSF21 | | | | | |
| Raw score | 8.58 (8.54 to 8.62) | 8.57 (8.53 to 8.61) | 0.01 (-0.05 to 0.06) | 0.781 | 0.998 |
| z-score | -0.14 (-0.35 to 0.07) | -0.18 (-0.42 to 0.05) | 0.04 (-0.27 to 0.36) | | |
| TNR | | | | | |
| Raw score | 4.97 (4.9 to 5.04) | 4.99 (4.91 to 5.07) | -0.02 (-0.12 to 0.09) | 0.748 | 0.998 |
| z-score | 0 (-0.19 to 0.19) | 0.05 (-0.17 to 0.26) | -0.05 (-0.33 to 0.24) | | |
| UNC5C | | | | | |
| Raw score | 5.05 (5 to 5.11) | 5.07 (5.01 to 5.14) | -0.02 (-0.1 to 0.06) | 0.651 | 0.998 |
| z-score | -0.12 (-0.28 to 0.03) | -0.07 (-0.24 to 0.1) | -0.05 (-0.29 to 0.18) | | |
| VWC2 | | | | | |
| Raw score | 6.26 (6.18 to 6.33) | 6.31 (6.23 to 6.4) | -0.06 (-0.17 to 0.06) | 0.306 | 0.935 |
| z-score | -0.05 (-0.26 to 0.16) | 0.11 (-0.12 to 0.34) | -0.16 (-0.48 to 0.15) | | |
| WFIKKN1 | | | | | |
| Raw score | 3.73 (3.68 to 3.79) | 3.79 (3.73 to 3.85) | -0.06 (-0.14 to 0.02) | 0.153 | 0.935 |
| z-score | -0.06 (-0.22 to 0.09) | 0.11 (-0.07 to 0.28) | -0.17 (-0.41 to 0.06) | | |

z Score values indicate how many standard deviations have the post-intervention values changed with respect to the baseline mean and standard deviation. E.g., a 0.50 z Score means that the mean value at post-intervention is 0.50 standard deviations higher than the mean value at baseline, indicating a positive change, with negative values indicating the opposite. Values are expressed as mean (95% CI). Analyses were adjusted for baseline values. N=87 participants. Raw scores values are expressed in Normalized Protein eXpression (NPX) units. This means that a high NPX value equals a high protein concentration. Because NPX is in a log2 scale, a 1 NPX difference means a doubling of protein concentration. If needed NPX values can be converted into linear scale: 2NPX= linear NPX. Abbreviations: $p_{FDR}$= adjusted for multiple comparisons using false discovery rate.
